# From Tribal Polarization to Socio-Economic Disparities: Exploring the Landscape of Vaccine Hesitancy on Twitter

**DOI:** 10.1101/2023.07.24.23293093

**Authors:** Huzeyfe Ayaz, Muhammed Hasan Celik, Huseyin Zeyd Koytak, Ibrahim Emre Yanik

## Abstract

This study analyzed Twitter posts related to vaccine hesitancy and its association with socio-economic variables in the US at the state level. The unique socio-economic characteristics of US states, such as education, race, or income, are significantly associated with attitudes toward vaccination. Our results indicate that vaccine hesitancy is a multifaceted phenomenon shaped by a complex interplay of factors. Furthermore, the research identifies two distinct sets of justifications for vaccine hesitancy. The first set pertains to political concerns, including constitutional rights and conspiracy theories. The second pertains to medical concerns about vaccine safety and efficacy. However, vaccine-hesitant Twitter users pragmatically use broad categories of justification for their beliefs. This behavior may suggest that vaccine hesitancy is influenced by political beliefs, unconscious emotions, and gut-level instinct. Our findings have further implications for the critical role of trust in shaping attitudes toward vaccination and the need for tailored communication strategies to restore faith in marginalized communities.

## 1 Introduction

Skepticism about the safety and benefits of vaccines is linked to low trust in institutions [1, 2]. Vaccine hesitancy, along with climate change denial, is part of a larger trend of mistrust in scientific expertise and a decline in trust in public institutions [3, 4, 5]. To combat the “infodemic,” the World Health Organization (WHO) has worked with major social media platforms to redirect internet users to reliable websites when searching for information related to COVID-19 [6]. Restoring confidence in scientific institutions is therefore an essential crucial democratic task for policymakers. This article endeavors to furnish those in positions of responsibility with a comprehensive understanding, while acknowledging the intricate nature of vaccine hesitancy and cautioning against the temptation of oversimplifying the issue by attributing it to a single social parameter, a mistake that many prominent media outlets have made. [7, 8, 9, 10].

Although there is no single universal determinant for vaccine hesitancy, a growing body of literature has identified factors that influence vaccine acceptance [11]. The Strategic Advisory Group of Experts (SAGE) on Immunization has categorized these determinants into three main domains, providing a framework for exploring the complex and multifaceted nature of vaccine hesitancy [12]. Addressing the contextual, individual/social group, and vaccine-specific determinants of vaccine hesitancy can help to overcome mistrust in vaccines and improve population health outcomes. Factors affecting vaccine acceptance are distinguished at different levels in scientific literature [13]. Our research focuses on social determinants of vaccine hesitancy at the population level, rather than on the socio-cultural context that influences individual decision-making.

A substantial body of literature has emerged on vaccine hesitancy to understand the factors that influence the uptake of public health interventions. Researchers have studied vaccine hesitancy to identify the pathways and mediators that contribute to a lack of public consent and resistance to various vaccines [11, 14, 15, 16, 17]. A key finding has been that vaccine hesitancy takes various forms and manifests differently [18], and that these self-destructive attitudes are unequally distributed among different segments of society [19] and across nations [20], exacerbating existing health disparities [21].

The attitudes toward vaccines are diverse and constantly evolving, making it important to understand their complexity [22, 23]. The views on vaccination have been found to be influenced by various sources of information, including the growing influence of digital public spheres [24, 25, 26, 27, 28, 29, 30]. Recent research has highlighted the significance of gathering and analyzing data from social media platforms in order to track the rapidly changing trends [29, 31], polarizations [32, 29], discourses [33, 34, 35], and sentiments [36, 37, 38] related to vaccine attitudes.

Recent advancements in computational social research have opened up new avenues for social research, particularly in analyzing the competing discourses in digital spheres. Social media data has been used to analyze societal attitudes toward critical social problems, such as immigration, public health, and extremism, due to its cost-effectiveness and ability to eliminate response biases [39]. In this study, we focused on analyzing vaccine hesitancy on Twitter, which offers easy access to large amounts of relevant content from millions of users through text mining techniques. Twitter provides several advantages as a data source in social research compared to conventional surveys. Firstly, it allowed us to analyze attitudes toward vaccines at a relatively low cost. Secondly, computational tools helped us uncover hidden patterns in a large number of Twitter messages with minimal human intervention. Unsupervised textual clustering methods provided an insider’s perspective on vaccine hesitancy while minimizing potential biases that could arise from the researchers themselves.

Our research demonstrates the complexity of vaccine-hesitant attitudes and the diversity of vaccine-hesitant populations. Our findings indicate that vaccine hesitancy is a multifaceted phenomenon, and the vast majority of vaccine-hesitant Twitter users pragmatically use different grounds to justify their stance. Additionally, we found that vaccine-hesitant content on Twitter comes from diverse geographical locations. While socio-economic factors do play a role in vaccine hesitancy at the state level in the U.S., our study suggests that attributing vaccine hesitancy solely to a single social factor, such as education, income, race, or voting behavior, is not appropriate. It is crucial to exercise caution when interpreting media reports and correlational studies that emphasize a single social determinant driving vaccine hesitancy. Our study provides new insights that can help to deepen our understanding of the underlying factors contributing to tribal polarization in the U.S. and revisit the relationships between vaccine hesitancy and social factors to improve public health outcomes.

## 2 Results

The study analyzed tweets from the first two years of the COVID-19 pandemic to gain insights into the online public discourse surrounding vaccination on Twitter. Tweets in the dataset were categorized based on their vaccination attitude, and the findings were presented in three parts: temporal analysis, spatial analysis, and topic modeling. The temporal analysis investigated the stability or variability of attitudes over time. The spatial comparison of geolocated tweets with socio-economic parameters revealed factors associated with vaccine hesitancy. Finally, the study used topic modeling of tweets to explore the most common justifications for vaccine hesitancy.

We utilized the Twitter Search v2 API (Methods) to fetch all English tweets containing the keywords ’vaccine’ and ‘vaccination’ from 2020 to 2022. Our resulting dataset comprises approximately 53 million tweets from around 8.2 million users, with roughly 584 thousand geolocated tweets for spatial analysis (Fig. 1.a). The vast majority of tweets within our dataset are representative of the United States. During the Eastern Standard Time (EST) daytime hours, we observed a peak in the number of tweets (Fig. S1), and a majority of geolocated tweets originated from the United States (Table S1). To eliminate any possible bias from non-US tweets, we further subset the dataset to solely include geolocated tweets originating from the United States for spatial analysis. We utilized the COVID-Twitter-BERT model [40] to obtain embedding vectors, which are numerical representations of tweets that capture their semantic meaning [41]. These embeddings facilitated downstream language tasks, such as classifying vaccination attitudes and conducting topic modeling.

**Figure 1.**
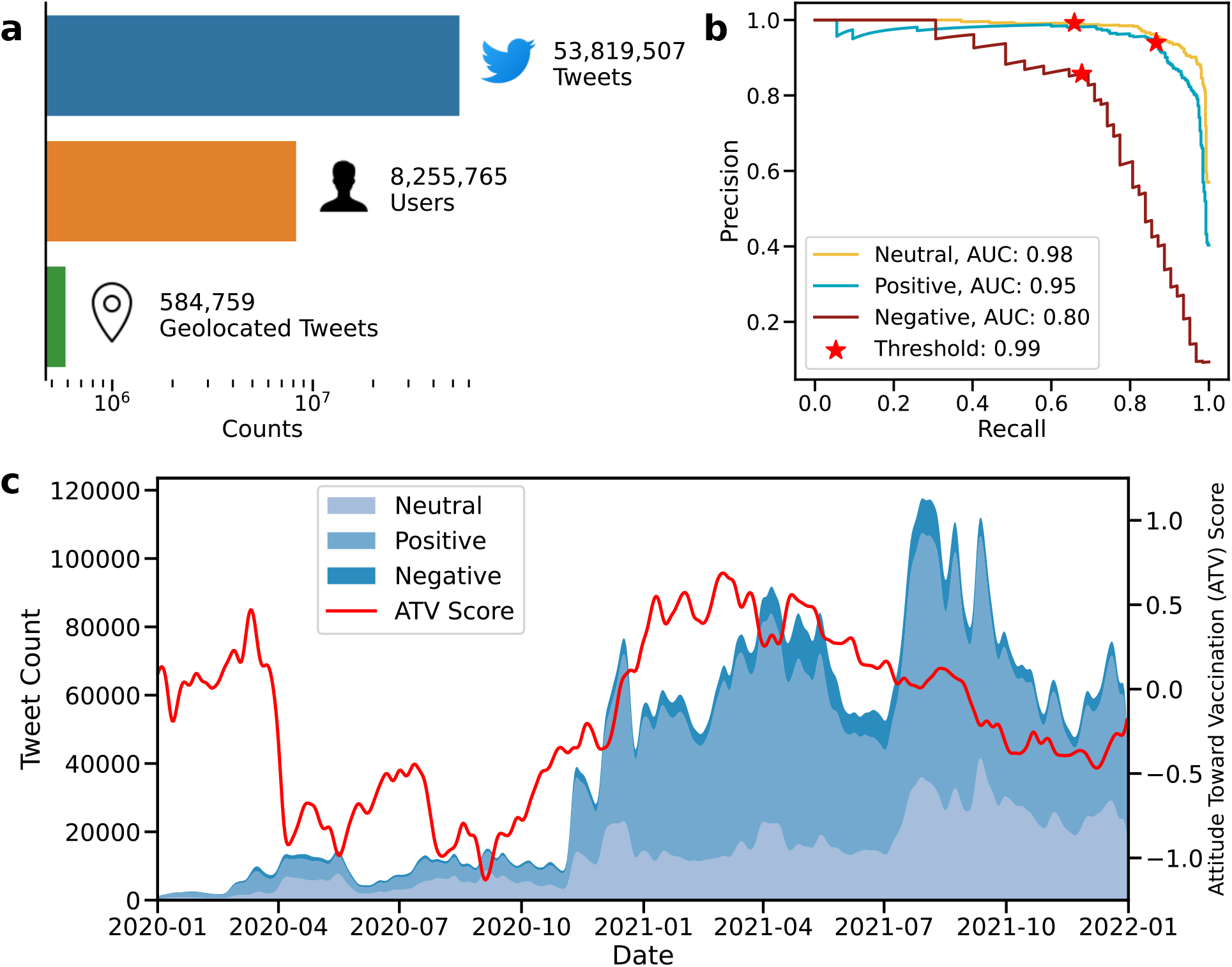
Dataset, attitude prediction. **(a)** Descriptive statistics of the Twitter vaccination dataset, such as the number of tweets, geolocated tweets, and users. **(b)** The precision-recall curve and AUC scores of the model for vaccination attitude classification. The red star indicates the threshold used in this study. **(c)** Temporal distribution of the number of neutral, positive, and negative tweets, and trend of the attitudes toward vaccination (ATV) scores.

A classification model was trained to predict attitudes toward vaccination by fine-tuning the BERT model on the previously annotated vaccination sentiment dataset [42] (Methods). The model achieved high performance, with PR-AUC scores of 0.98 for neutral, 0.95 for positive, and 0.8 for negative classes on the test set (Fig. 1.b). Neutral tweets were filtered out, and only positive and negative tweets with a predicted class probability above 99% were used to ensure high precision (Fig. S2). At the proposed threshold (indicated with a red star in Fig. 1.b), the model achieved a precision of 94% at 87% recall for the positive class and an 86% precision at 68% recall for the negative class (Table S2). Notably, the classification performance reported in this study outperforms the Fasttext-based approach of Muller et al. (2019), which was reported as 77% for precision and 77% for recall (Table S3).

The study developed attitudes toward vaccination (ATV) score, which summarizes the proportion of positive and negative tweets in terms of log odds ratio by their location or the specific time period they were posted. The ATV score was calculated per day using all tweets and by the US state using only geolocated tweets. The ATV scores of geolocated tweets are representative of all tweets based on monthly ATV score comparison (Spearman’s rank correlation coefficient = 96%, *P* = 6 *∗* 10^−5^), as shown in Fig. S3.a,b. The study also investigated the relationship between the vaccination rate of each US state by December 31st, 2021 [43] and state-level ATV scores, finding a high correlation (Spearman’s rank correlation coefficient = 53%, *P* = 6 *∗* 10^−5^) that indicates the proposed ATV score is representative of the vaccination behavior of a broader US population beyond the Twittersphere (Fig. S4.a,b).

In order to understand the stability and variability of attitudes toward vaccination, we investigated the timeline of two years (Fig. 1.c). The attitude was defined as stable if the vaccination attitude did not change over time, and as variable if the perspective on vaccination differed over time. While positive tweets about vaccines predominated over negative tweets during the entire timeline, the relative ratio of positive to negative tweets fluctuated. The first quarter of 2020 had a small fraction of all tweets, comprising less than 0.5% of the entire dataset, which could be explained by low COVID-19 death numbers in the USA. However, the ATV score decreased sharply with rapidly increasing COVID-19 death numbers in late March and stayed relatively low until November 2020. This period also had a high unemployment rate, although it peaked in early May at 15.8% [44] (Fig. S5). During 2021, vaccination had greater online public attention, and the aggregate attitudes toward vaccination were elevated in any quarter of 2021 compared to 2020 (Fig. S6), parallel to the successful vaccine trials and the increasing vaccination rate. However, the variability of the aggregate vaccination attitudes may not imply a shift in individuals’ views toward vaccination because the composition of users differs over time.

To address this, we conducted a user-level analysis by tracking the vaccination attitudes of users who posted multiple tweets across the studied timeline. We found that 31% of users had more than one tweet in the dataset, and most users were clearly polarized into positive and negative attitude camps by predominantly tweeting with a single attitude (Fig. 2.b, Fig. S7). There was no apparent difference between these groups based on the number of tweets they posted (Fig. S8.a,b). Also, there were only a few outliers with more than thousands of tweets, indicating that few users do not disproportionately influence our analysis (Fig. 2.a). The percentage of positive tweet distributions of users before and after November 1st, 2020 is shown in Fig. 2.b, and there is a statistically significant shift to a more positive opinion. Specifically, users are 11% more likely to post a tweet with a positive attitude after November 1st, 2020, compared to their past selves. We repeated this analysis for all the dates in our dataset and observed that attitudes toward users decreased in the first five months of 2020 and increased afterward, except in the last four months of 2021, when there was no significant change (Fig. 2.c, Fig. S9.a,b).

**Figure 2.**
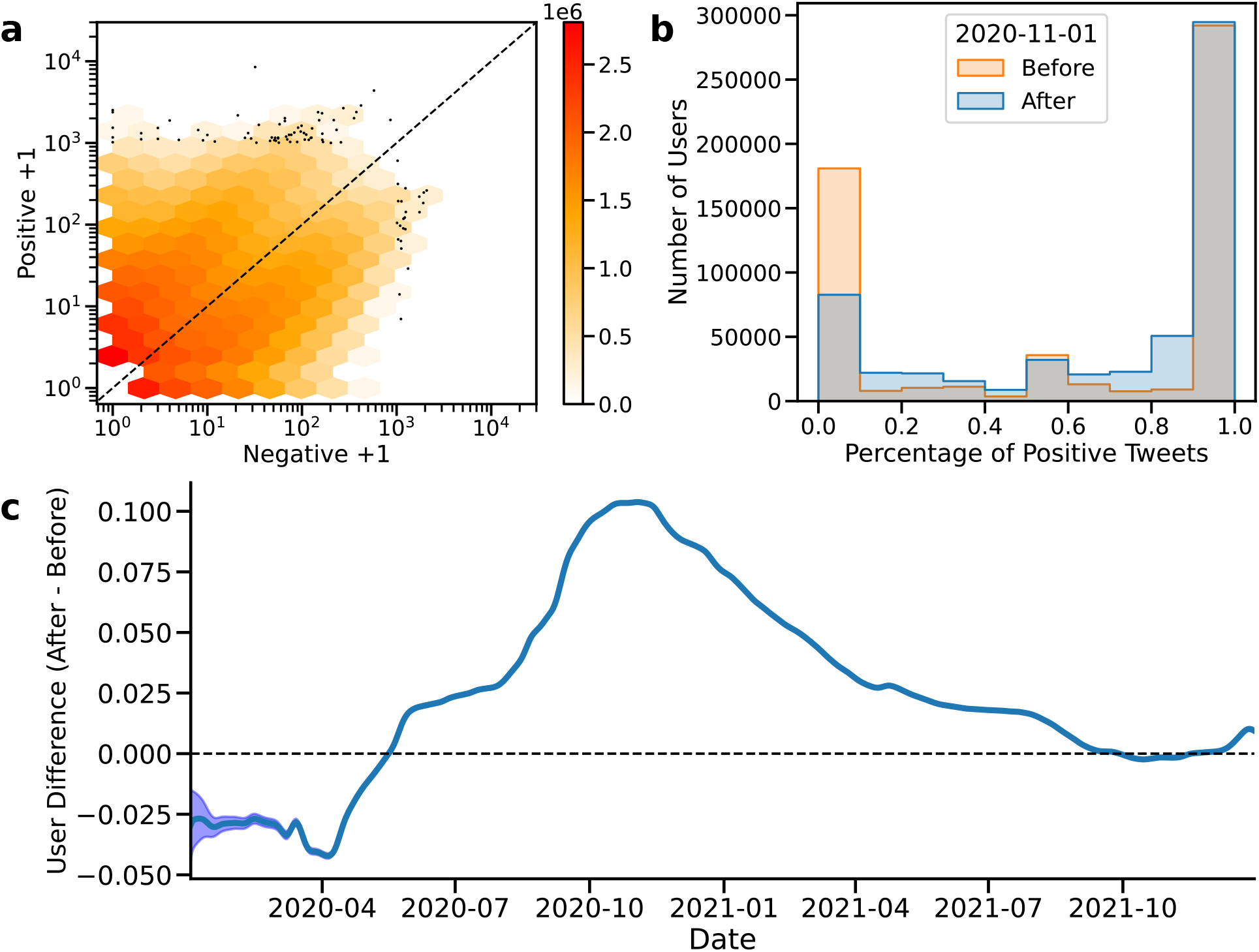
Temporal change of attitudes. **(a)** The number of tweets posted by users **(b)** The percentage of positive tweets of users with at least two tweets before and after November 1st. **(c)** The trend of vaccination attitudes at the user level.

We conducted an even simpler analysis with the users who posted two tweets (n=906,430). The comparison of their first tweet against their later tweet shows a similar trend to a more positive attitude, regardless of the date. Only 12% of users with a positive first tweet later posted a negative second tweet, while 40% of users with a negative first tweet posted a positive second tweet (Table S4). Although the majority of users did not change their attitudes toward vaccination, there is a significant minority of negative users who have switched to a more positive attitude. Overall, both aggregate and user-level analysis indicate that a significant minority of users have more positive attitudes; yet the vast majority of users retained that vaccination attitude.

To investigate the socio-economic factors associated with vaccine hesitancy, we calculated the ATV score by the US states based on geolocated tweets and identified distinct regions based on attitude (as shown in Fig. 3.a). Using Fisher’s exact test, we found that 25 out of 50 states have a significant pro-vaccine stance, while the other 25 have an anti-vaccine stance (Fig. 3.b and Table S5). We collected 10 socio-economic parameters for each state (Methods) and found that 8 out of 10 socio-economic parameters are significantly correlated with vaccination attitude based on univariate correlation analysis (Fig. S10.a-j). For instance, there is a negative correlation of −49% (*p* = 3 *∗* 10^−4^) between ATV score and having less than a high school degree (Fig. S10.b), while there is a positive correlation of 45% (*p* = 10^−3^) between voting for Trump in 2020 and ATV (Fig. S10.h). However, we also found that the higher rate of cat ownership compared to the rate of dog ownership has a 46% Spearman’s rank correlation coefficient (Fig. S10.j), indicating the limitation of the univariate analysis, which may not accurately represent the underlying socio-economic factors related to vaccination attitude. It is important to note that socio-economic factors are not easily separable but rather interconnected (as depicted in Fig. 3.c), and therefore, multicollinearity between socio-economic parameters may lead to spurious correlations.

**Figure 3.**
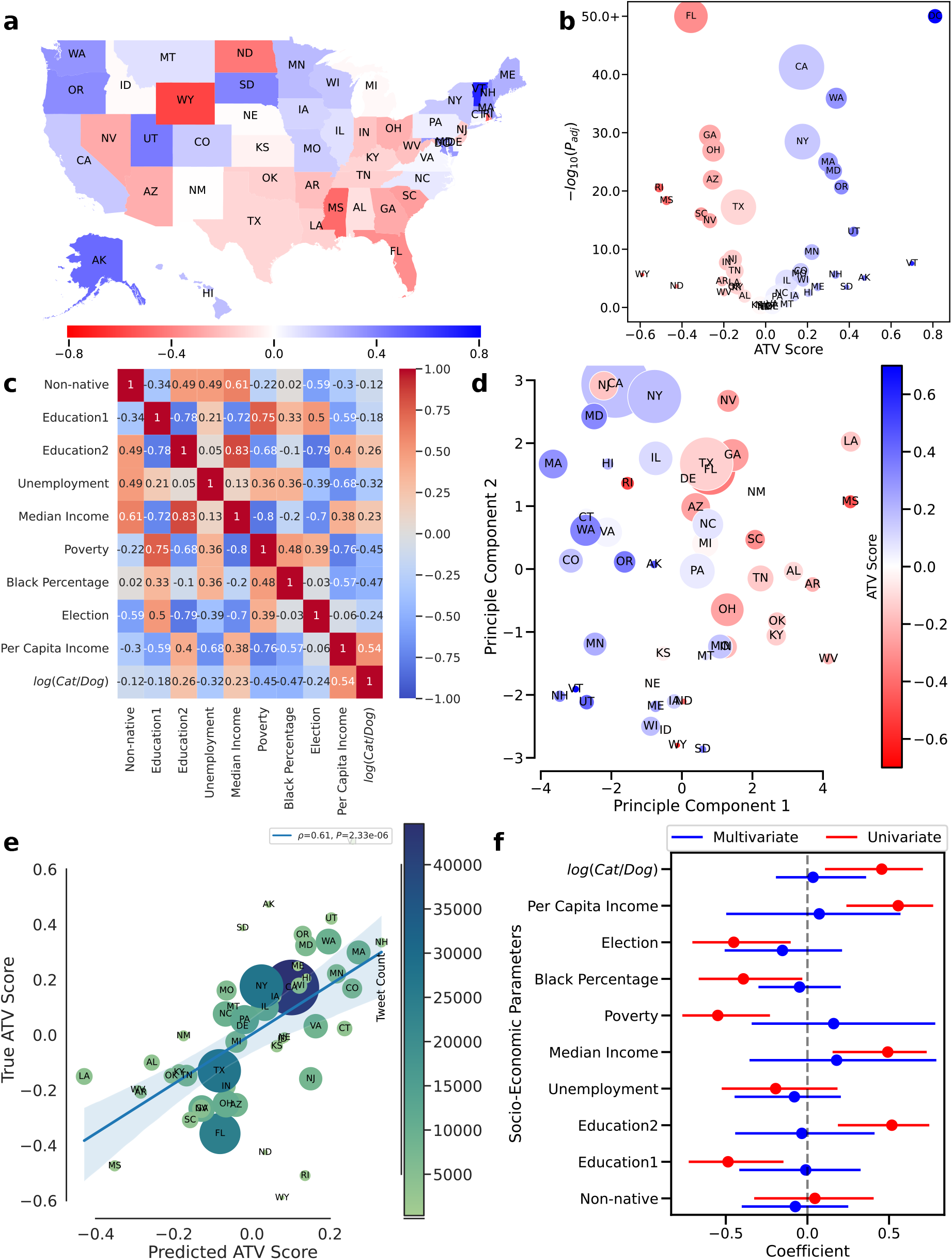
State-level socio-economic parameters and attitudes toward vaccination. **(a)** U.S. states colored by ATV score. **(b)** Volcano plot of ATV score and statistical significance of vaccination attitude of each state **(c)** Cross-correlation of socio-economic parameters with Spearman’s method **(d)** The scatterplot of the first two principal components of the US states, which is colored by ATV-score. **(e)** True and predicted ATV scores based on PLS regression **(f)** Correlation coefficients of socio-economic parameters based on univariate (Spearman’s correlation coefficient) and multivariate (coefficients of PLS regression) analysis with error bars.

PCA (principal component analysis) was used to identify orthogonal latent factors from the socio-economic parameters. The first two principal components, which represent the characteristics of each state, were plotted in Fig. 3.d (Fig. S11, Fig. S12, Table S6). Each state is then colored according to its ATV score. Remarkably, even though the PCA analysis does not utilize the ATV score as a feature, the states are evidently separated based on their attitudes toward vaccination, primarily along the first principal component. This finding strongly suggests that unique state-level characteristics are associated with vaccination attitudes.

We developed a partial-linear (PLS) regression model to predict the ATV score of each state based on the socio-economic parameters (Methods). The PLR regression, similar to PCA, identifies orthogonal latent factors for features and correlates those factors with the ATV score, which overcomes the collinearity that conventional regression models suffer from. The predicted ATV scores based on socio-economic parameters using the PLS model have a Spearman correlation coefficient of 61% (*p* = 2.33*e* − 06) against ground-truth ATV scores based on the one-leave-out test set (Fig. 3.e). This result indicates that socio-economic features are highly predictive of ATV scores. We conducted a bootstrapping analysis to investigate specific socio-economic parameters linked to vaccine hesitancy when other parameters remained constant (Methods). Confidence intervals of coefficients obtained from bootstrapping analysis were adjusted for a false discovery rate of 5% to avoid multiple comparisons (Fig. 3.f). The analysis reveals that none of the socio-economic features is significant alone when controlled for other socio-economic parameters.

Overall, these results suggest that states have unique characteristics defined by socio-economic parameters that separate them into two camps and are highly predictive of attitudes toward vaccination. Yet, it is not possible to obtain the definitive link between vaccine attitude and a specific socio-economic parameter through state-level analysis without further research.

The common justifications for vaccine hesitancy were obtained by performing contextual topics modeling analysis (Methods) on the tweets with negative attitudes. As a result, seven main themes for vaccine hesitancy were discovered (Table 1, Fig. S14, Table S7). Similar themes have been reported in the childhood vaccination literature. For instance, Majid and Ahmad (2020) examined 34 qualitative studies to clarify parents’ reasons for rejecting or delaying vaccines [45]. Although their examination did not cover the studies on COVID-19 vaccine hesitancy, the fear of side effects and skepticism regarding vaccine effectiveness were prevalent among parents who rejected vaccines, which is consistent with what we found in our topics. Additionally, some of the studies they examined highlighted that distrust in health system players and mandatory vaccine policies were reasons why parents rejected vaccines for their kids. In our study, we encountered a similar stance on the alleged conflict of interest in the health system and categorized it as politics and conspiracy theories. Similarly, mandatory vaccine policies are a source of frustration for many vaccine-hesitant Twitter users in our research. This demonstrates parallelism between the ways in which people justify their vaccine-hesitant attitudes in the face of the COVID-19 pandemic and previous waves of anti-vaxxer movements.

**Table 1:**
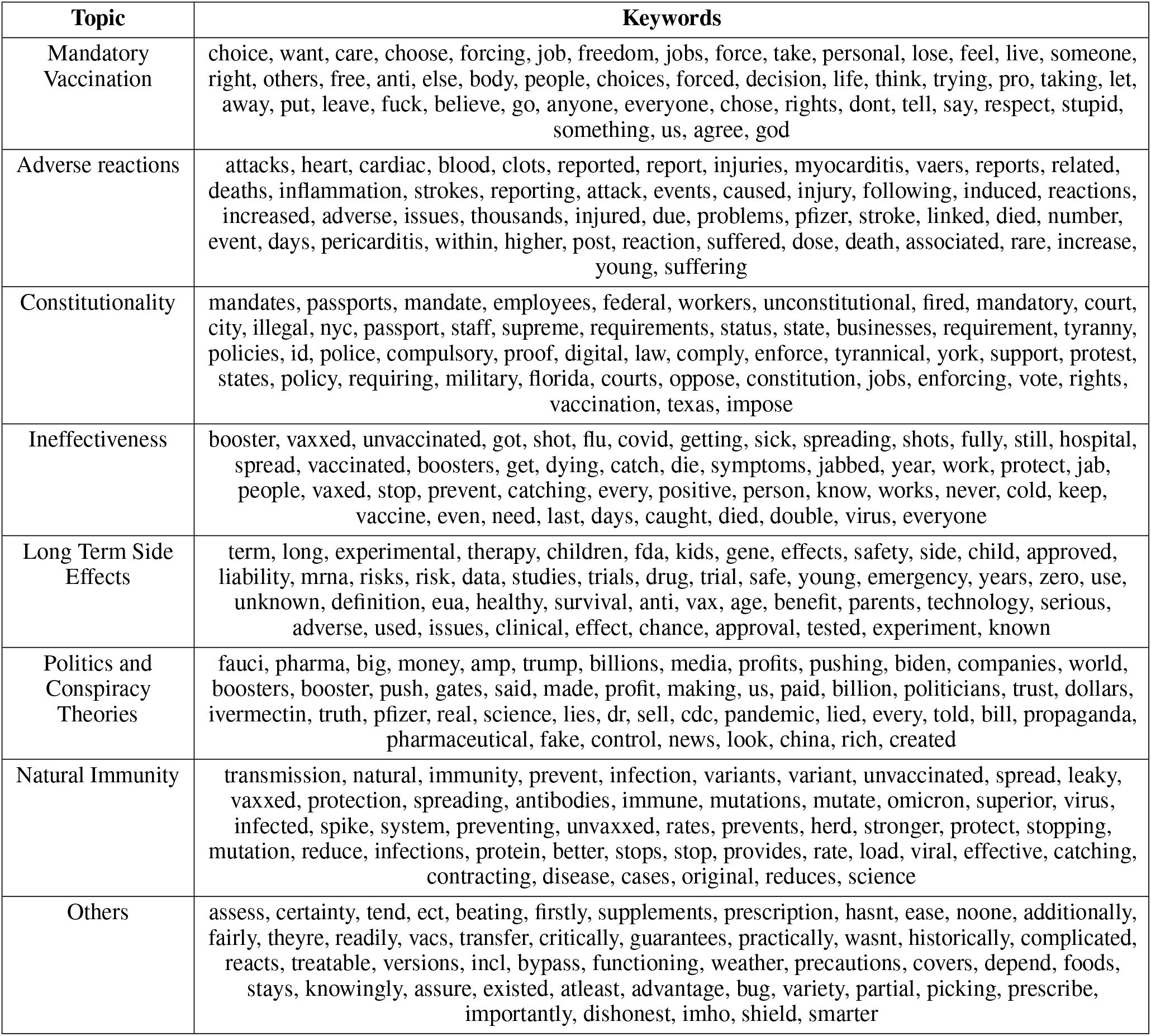
Themes identified by topic modeling and top keywords describing each theme.

Using a topic modeling technique, we have identified eight themes related to vaccine hesitancy in our dataset. The first theme covers discussions on mandatory vaccinations and features social media conversations about vaccine administration practices. Many vaccine hesitant Twitter users framed mandatory vaccination as a violation of their individual rights. Another theme pertains to the constitutionality of vaccine mandates, such as vaccine passports and vaccination requirements for state and federal workers. Our insights from these two themes echo recent research in the history of vaccination, which has documented how activists in US history opposed vaccination on the basis of rights, freedom, and liberty, even when vaccination was not compulsory. [46] This work highlights the ongoing tension between scientific expertise and civic freedom to choose one’s own medical practices. In social media, we see similar traces of this tension that led to the Jacobson v. Massachusetts case in 1905. [46, 47]

The Twitter discussions focus heavily on the health concerns of users, with several themes emerging. One theme in this category is the alleged adverse reactions of vaccines. Another related theme focuses on the long-term side effects of COVID-19 vaccines. Vaccine hesitant tweets on this topic discuss potential long-term side effects, which may arise from the novel mRNA method or the risks associated with the expedited vaccine approval process. “Ineffectiveness” is another health-related topic that covers skepticism over the efficiency of vaccines or their total denial. A systematic review found that safety and side effects were the two most commonly reported factors influencing COVID-19 vaccine hesitancy in various studies [48]. Another meta-analysis concluded that stronger beliefs in the unsafety of vaccines are among the predictors of vaccine hesitancy globally [49]. The belief that COVID-19 vaccines are unsafe or ineffective is the factor that has been supported by the largest number of studies in high-income countries, while concerns about the rapid development of vaccines follow them on the list [50].

A final set of health-related vaccine hesitancy themes involves social media content that either partially or completely denies the benefits of vaccines. The “natural immunity” theme suggests that natural immunity is superior to vaccines in terms of protection against the virus. Politics and conspiracy theories are closely linked to COVID-19 denialism in health-related tweets. Twitter users discussed the alleged conflict of interest in the production of scientific knowledge and its dissemination by the mass media in line with the political frame of their choice. The relationship between belief in conspiracy theories and vaccine hesitancy has been extensively studied [51, 52]. However, the contribution of conspiracy theories to vaccine hesitancy has been overstated without solid empirical evidence. In fact, an independent research center has found that only a few people were responsible for the vast majority of misleading content about vaccines on social media [53]. In terms of substance, the Big Pharma conspiracy theory predates the COVID-19 pandemic and uses cui bono reasoning to identify a small number of elites who benefit from either deliberately manufacturing or not fully curing diseases [54].

The extracted substantial themes, including the seven main themes and the “others” category, reveal the contours of the vaccine hesitant debate on Twitter. Conspiracy theories with political implications represent only one aspect of vaccine hesitant content; there is much more to it. Vaccine administration and health concerns are the two primary branches of vaccine hesitancy in social media discussions. Both of these strands encompass a wide range of themes, from moderate skepticism to extreme denialism. Our model’s vaccine hesitant topics are consistent with previous studies on vaccine hesitancy, which have also identified concerns about side effects, lack of trust, and belief in the ineffectiveness of vaccines as subthemes [55].

Earlier in our study, we illustrated that vaccine hesitancy takes on many shades and facets. To gain a better understanding of how vaccine-hesitant individuals rationalize their opposition to COVID-19 vaccines, we explored the extent to which they indiscriminately adopt arguments from other prevalent themes of vaccine hesitancy. Specifically, we examined whether users were solely focused on one particular topic or if their concerns spanned across multiple topics. To achieve this, we categorized users by topics if they had posted at least one tweet on the subject. Then, we measured the topic distribution of their remaining tweets, as illustrated in Fig. 4. We observed that users in our dataset did not exhibit a consistent pattern of opposition based on a single theme within vaccine hesitancy topics. Instead, they demonstrated a high degree of variability across topics, not limiting themselves to a specific theme. These individuals employed a broad range of topics to rationalize their vaccine hesitancy. Fig. 4 portrays the co-occurrence of the same users across various vaccine hesitant themes. For instance, a user who posted their first tweet on adverse reactions might subsequently employ an argument from the conspiracy theory theme to bolster their stance. Overall, the desultory use of arguments from a wide range of topics may imply that users are potentially motivated by deeper prejudices, and they utilize any available justification in their Twitter posts.

**Figure 4.**
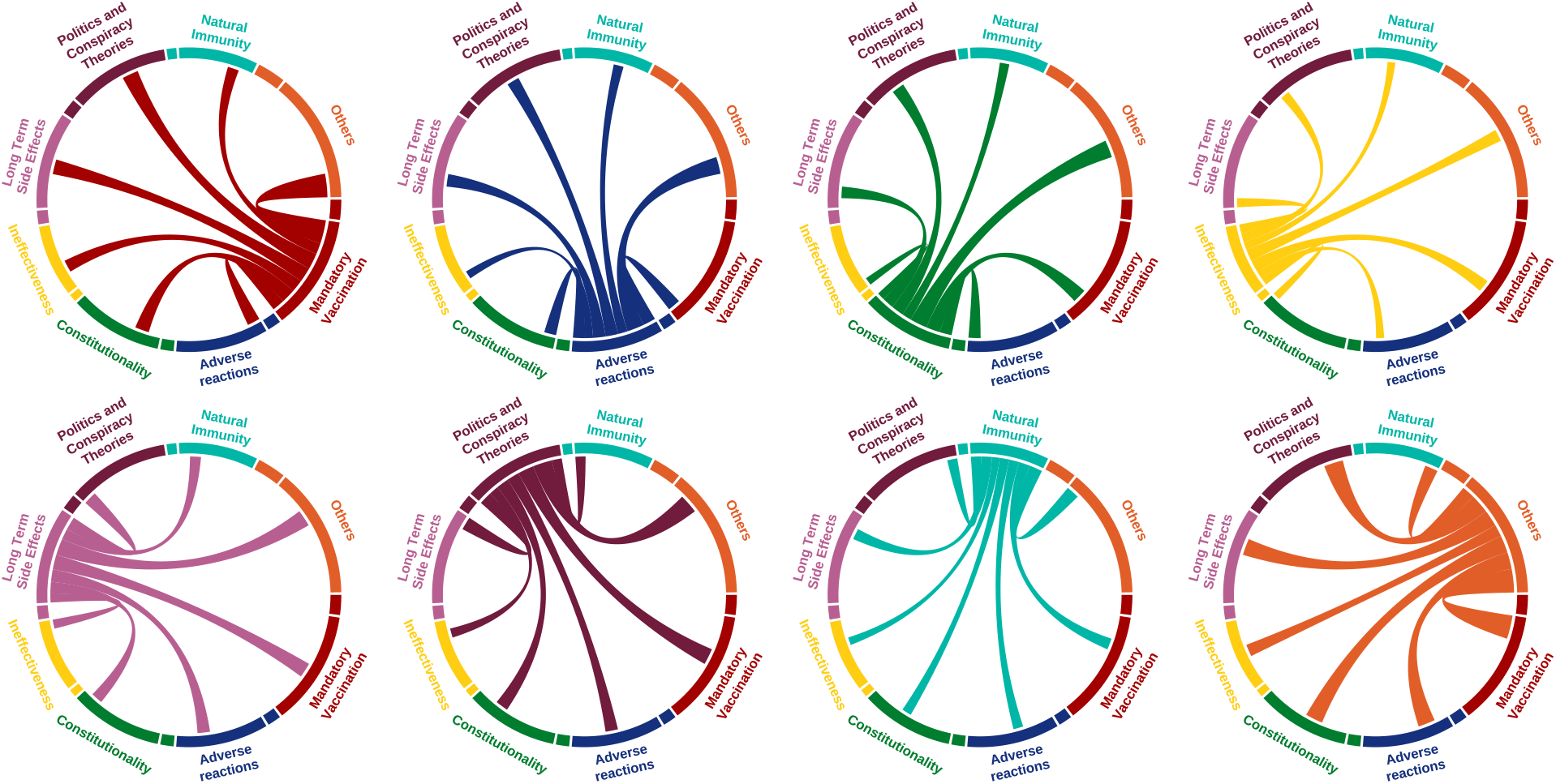
The topic distribution of Twitter posts of users who posted multiple tweets. Each diagram represents a group of users interested in a specific topic and illustrates how their other tweets are distributed.

## 3 Discussion

In this study, we investigate skepticism towards vaccination by analyzing public discussions on Twitter using state-of-the-art natural language processing techniques. Our analysis sheds light on the polarization of online communities regarding COVID-19 vaccines.

The temporal aspect of our study design reveals the rigidity of attitudes towards vaccines. Despite the scientific advancements and approval of the first COVID-19 vaccines, only a small minority of Twitter users have shown a positive change in their attitudes towards vaccines. In addition to the composition effect, we also observe a slight optimism regarding the pandemic. However, negative attitudes towards vaccination have mostly persisted within the online community.

To delve deeper into the attributes of the polarized online communities, we used a novel methodology by merging geolocated online content with the conventional socio-economic variables based on the location of Twitter messages. Our association analysis between socio-economic variables and vaccine hesitancy demonstrated that socio-economic parameters can predict the degree of vaccine hesitancy in US states. The residents of those states suffer from higher rates of unemployment, lower median household income, and poorer educational outcomes. However, the vast disparities in vaccine hesitancy among US states cannot be solely attributed to a single social parameter, such as election results, education, race, or income. Vaccination hesitant online communities have unique characteristics defined by a complex amalgamation of socio-economic parameters.

To gain further insights into discourses of the vaccine-hesitant community, we conducted a textual analysis and identified various themes revolving around two main concerns, political and medical. Political concerns branch out to two different themes: constitutional issues related to vaccine mandates and conspiracy theories. Likewise, medical concerns include two dissimilar themes: medical side effects and the denial of their effectiveness. These concerns are similar to the reasons to refuse vaccines identified by a systematic review of earlier work as (1) medical safety of COVID vaccines, (2) the inefficiency of vaccines, and (3) belief in natural immunity [56]. Yet, Twitter users who posted multiple times are not fixated on a single issue, rather they pragmatically borrowed arguments from a wide range of vaccine hesitant themes and they often justified their stances against vaccines using multiple reasons.

Several studies have indicated that marginalized communities are less likely to trust institutions, including vaccines [57, 58]. For example, working-class Whites are one such group, often portrayed as former President Trump’s base [59]. They inhabit areas characterized by high levels of poverty and elevated rates of premature deaths due to gun violence, suicide, drug overdoses, and alcoholism, compared to the national average [60, 61]. The historical marginalization of those communities leads to a higher concern about government intrusion in their personal lives. As a result, those particular segments of society feel fear and anxiety in the face of strict vaccine mandates and protocols.

The moral foundations theory can illuminate our findings regarding the vaccination hesitant community [62]. The theory suggests that people primarily rely on moral intuitions shaped by socio-psychological factors to make political judgments, then they justify their judgments with strategic reasoning [62]. Some Twitter users’ attitudes toward vaccines may have been shaped by emotions and gut-level feelings, leading to distrust of institutions that underlies vaccine hesitancy. Our findings regarding the spurious relationship between a solitary socio-economic parameter and vaccine hesitancy as well as the indiscriminate use of different topics may support this idea. This distrust may stem from a heightened moral preference expressed through a series of ad-hoc hypotheses, as classified as diverse vaccine-hesitant themes in our study [63].

To address these concerns, a well-planned public communication strategy is necessary. Understanding the moral foundations of their attitude and establishing empathy should be the initial step. One approach to restore faith and rebuild trust in those communities would be to adopt a persuasive language that emphasizes the common good, with the support of local leaders. Without the support of their in-group, public health efforts may be seen as stigmatizing and compulsory measures by marginalized communities [58]. Communication strategies tailored to local contexts may improve to trust in science.

## 4 Methods

### 4.1 Fetching Tweets

In order to fetch tweets, we utilized searchtweets-v2 [64] and Tweepy [65] python packages. Tweets were fetched using the search endpoint of the Twitter-API (https://api.twitter.com/2/tweets/search/all). We fetched all tweets containing the keywords: “Vaccine” or “Vaccination”. We limited our analysis with those English tweets posted between 2020 and 2022. We excluded retweets, short, (less than 10 words) and duplicate tweets. Then we used the geo-location identifier of tweets to locate the state where tweets were posted from by using Tweepy library.

### 4.2 Dataset and Model Training For Vaccination Attitude Prediction

To identify individuals who express vaccine hesitancy, tweets were classified into one of three categories of “Positive”, “Negative”, or “Neutral” by using BERT-based natural language processing (NLP) models. BERT embeddings were extracted using Covid-Twitter-Bert-V2 [66] model from HuggingFace [67] platform. Then, the model was fine-tuned on the dataset by Pananos et. al. [42] to predict vaccination attitude. The dataset is consists of 27,906 tweets which were manually labeled by Amazon Mechanical Turk. The annotators of the crowdsourced dataset have a consensus for only 16,156 tweets. The dataset is partitioned into the train, validation, and test sets with proportions of 80%, 10%, and 10%, respectively. To create the validation and test sets, we only included tweets with consensus. The remaining tweets, with or without consensus, were used for fine-tuning with SimpleTransformers library [68]. During the fine-tuning, the model was trained for 20 epochs with the early stopping of 5 epochs. To optimize our model’s performance, we conducted hyperparameter tuning for several parameters. The key parameters that we hyperparameter-tuned were learning rate, epsilon for Adam optimization, and maximum sequence length. For the learning rate, we experimented with three values: 1e-3, 1e-4, and 1e-5. Similarly, we tried three values for epsilon, 1e-7, 1e-8, and 1e-9. Also, class weights were calculated based on the inverse proportion of class frequencies to ensure balanced training. The weights used were 1.0, 1.43, and 9.2 for Neutral, Positive, and Negative, respectively. The best score was obtained with the parameters of 1e-05 for learning rate, 1e-07 for epsilon, and 128 for maximum sequence length. The performance of our model was evaluated using the f1-macro metric using scikit-learn [69] during the hyperparameter tuning.

### 4.3 Prediction of Attitudes Toward Vaccination Score

We used the fine-tuned model to predict the label of all fetched tweets and only used high-confidence predictions of the model where the predicted class probability is above 99%. To analyze spatial and temporal dimensions of vaccination hesitancy, we defined the attitudes toward vaccination (ATV) score, which is log odds ratio (OR) positive *p* to negative *n* tweets for specific category *c* (place or date):

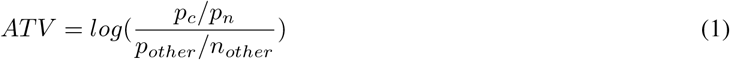

To obtain the statistical significance of attitudes toward vaccination scores by the US states, we also employed Fisher’s Exact Test [70] with categories of attitudes (positive and negative) and the state tweet was sent from. P-values of Fisher’s Exact test were corrected for multiple testing using Bonferroni correction method.

### 4.4 Socio-Economic Parameters and ATV Score

To investigate the relationship between vaccine hesitancy and socio-economic parameters, we used 2020 American Community Survey 1-Year Experimental Estimates data by the U.S. Census Bureau. The parameters included the percentage of Black population (Black percentage), the percentage of people living in poverty in the past 12 months (Poverty), inflation-adjusted median household income in the past 12 months (Median income), employment status for individuals aged 16 years and over (Unemployment), and citizenship status (Non-native). We also studied the educational attainment of the population aged 25 years and over, specifically the percentage of those with less than a high school degree (Education1) and those with more than a bachelor’s degree (Education2). Additionally, we examined the influence of presidential elections (Election) on attitudes toward vaccination using the results of a study [71]. We also included Social Capital (PSU-SC) index (Rupasingha et al. (2006 with updates) [72]) and Cat/Dog ownership ratio by the US states [73] as a dichotomous variable 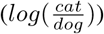.

We calculated Spearman’s rank correlation coefficient for each socio-economic parameter. To find the error bars that are corrected for multiple testing, we used the formula in equation 2:

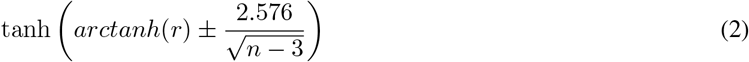

where *r* is the estimate of the correlation and n is the sample size. 2.576 is the adjusted z-score for a 99% confidence interval with the Bonferroni correction.

### 4.5 Partial Least Squares (PLS) Regression

For the multivariate analysis, we used Partial Least Squares (PLS) Regression [74] to avoid multicollinearity between the socio-economic parameters. Bootstrapping was conducted for 10,000 iterations to obtain error bars for regression coefficients with 99% confidence interval. In each bootstrapping iteration, the model was fitted on the random subset of states.

### 4.6 Contextualized Topic Modeling (CTM)

We utilized the Contextualized Topic Modeling (CTM) [75] library, which is a Variational AutoEncoder (VAE) [76] based deep learning model. CTM takes BERT embeddings as an input and predicts a bag of words (BoW) by sampling from the bottleneck layer. The bottleneck size is equal to the number of topics. This approach enabled us to assign a topic to each tweet with an unsupervised method. We conducted topic modeling with only negative tweets. NLTK [77] library is utilized to remove stop words. The number of topics, hidden layer sizes of the encoder and the decoder, and the dropout rate are hyperparameter tuned. We tuned topics between 5 to 10, unit size of the hidden layer of 200, 500, and 700, and dropout rates of 0.2, 0.5, and 0.8. All hyperparameter tuning experiments were done in 2 epochs. The best hyperparameters were chosen based on topic coherence and diversity metrics. We selected the topic number as 8, a hidden layer size as 200, and a dropout as 0.2. Topic coherence is the average normalized pointwise mutual information (NPMI) score between the top keywords in a topic. The formula for topic diversity is defined as follows:

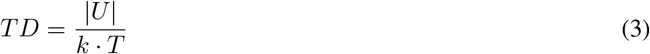

where TD is the topic diversity score, |U| is the number of unique words in the corpus, k is the number of top keywords of each topic, and T is the total number of topics in the model. Top keywords for each topic are generated with 20 steps of post-training sampling from the bottleneck layer. Keywords with high Pointwise Mutual Information (PMI) score (greater than 6) were merged into a phrase. We assigned tweets to topics when the topic probability was marginally greater (at least 12.5%) than other topic probabilities, and then the remaining unassigned tweets were filtered.

## 5 Data and Code Availability

The code used for this project is available on GitHub at https://github.com/twittersphere/twitter_vaccine_hesitancy. The Attitude and Contextualized Topic Modeling predictions generated in this study are available on Zenodo at https://doi.org/10.5281/zenodo.7876072.

## Data Availability

All data produced in the present study are available upon reasonable request to the authors.

