## Supplementary Material for "From Tribal Polarization to Socio-Economic Disparities: Exploring the Landscape of Vaccine Hesitancy on Twitter"

**Huzeyfe Ayaz**[*]
Department of Informatics
Technical University of Munich
Garching, Munich, Germany

**Muhammed Hasan Celik**[*†]
Department of Computer Science
Center for Complex Biological Systems
University of California Irvine
Irvine, CA, USA


**Huseyin Zeyd Koytak**[*†]
Department of Sociology
Syracuse University
Syracuse, NY, USA


**Ibrahim Emre Yanik**
Department of Sociology
Syracuse University
Syracuse, NY, USA

***Keywords*** Vaccine hesitancy · Trust in institutions · COVID-19 · political beliefs · Twitter

[*]All authors contributed equally
[†]Author to whom correspondence should be addressed

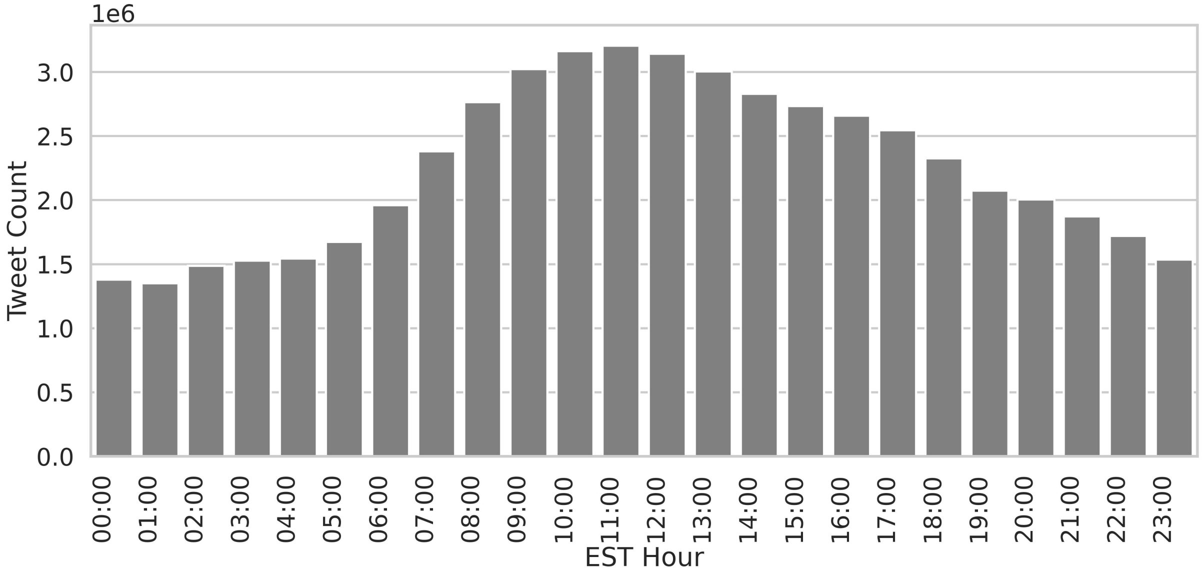

Figure 1: The hourly tweet count based on Eastern Standard Time (UTC-5). The results show that the number of tweets peaks during the daytime hours of EST.

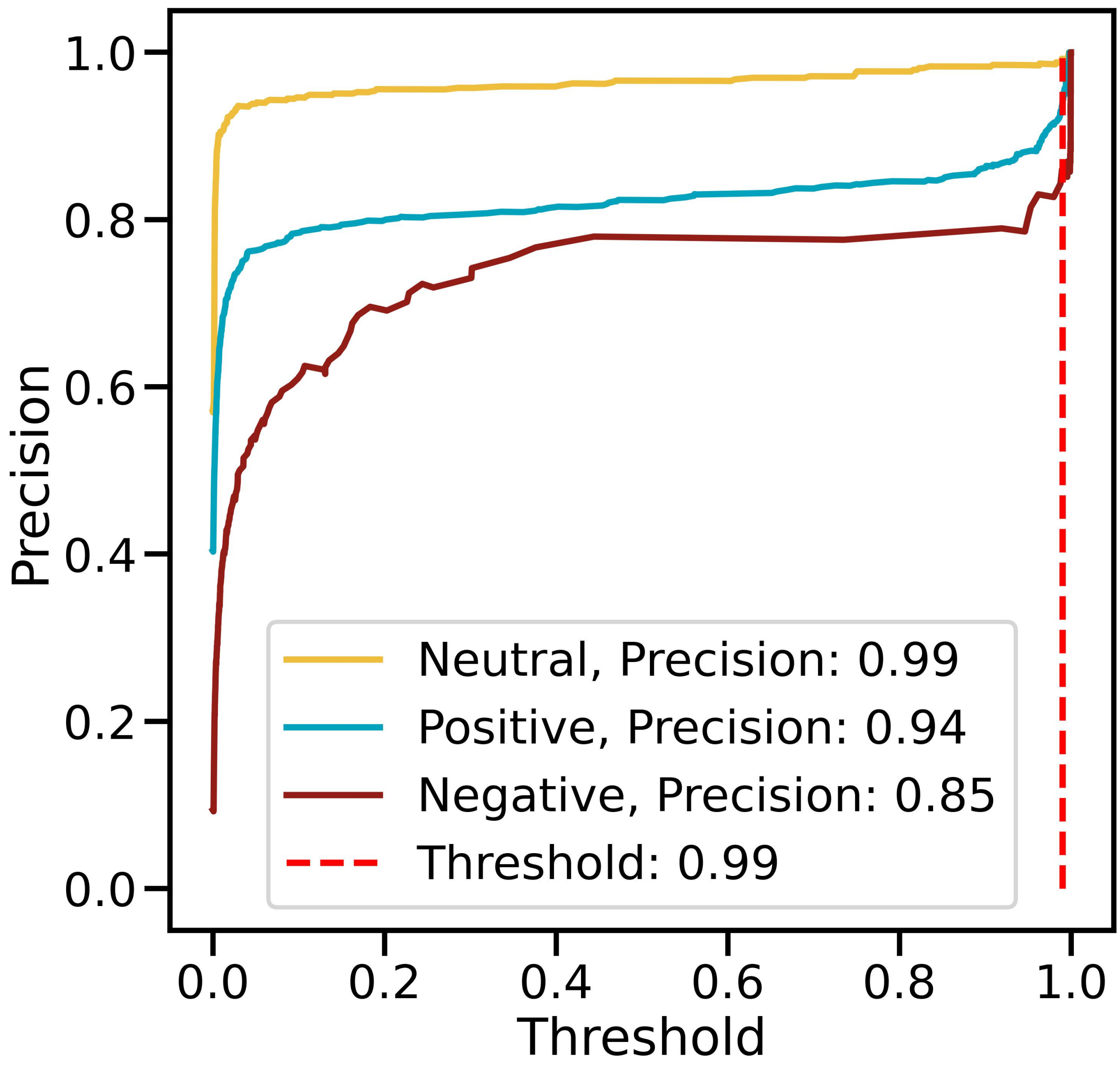

Figure 2: The precision-threshold curve for the proposed attitudes toward vaccination model illustrates the relationship between precision and threshold for classifying tweets as Neutral, Positive, or Negative. Precision is a measure of correctly classified samples out of all samples classified as a specific class, while the threshold is the minimum class probability required for a class assignment. The dashed red line represents a threshold of 0.99.

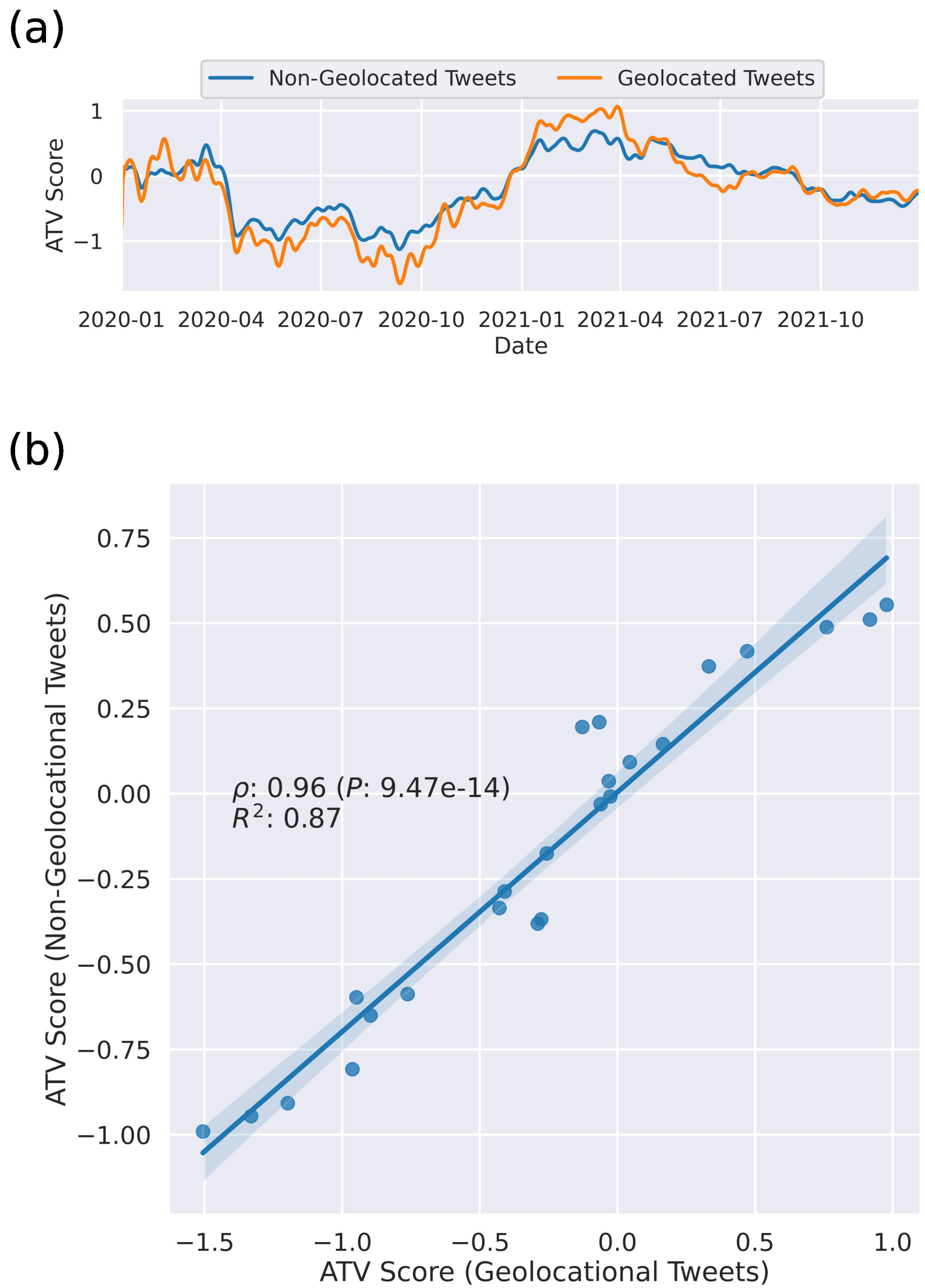

Figure 3: **(a)** This figure displays the daily ATV score for geolocated and non-geolocated tweets between 2020 and 2022. Geolocated and non-geolocated tweets follow the same trend. **(b)** Monthly ATV scores correlation between geolocated and non-geolocated tweets. There is a high correlation (R-squared of 87%) between ATV scores of geolocated and non-geolocated tweets.

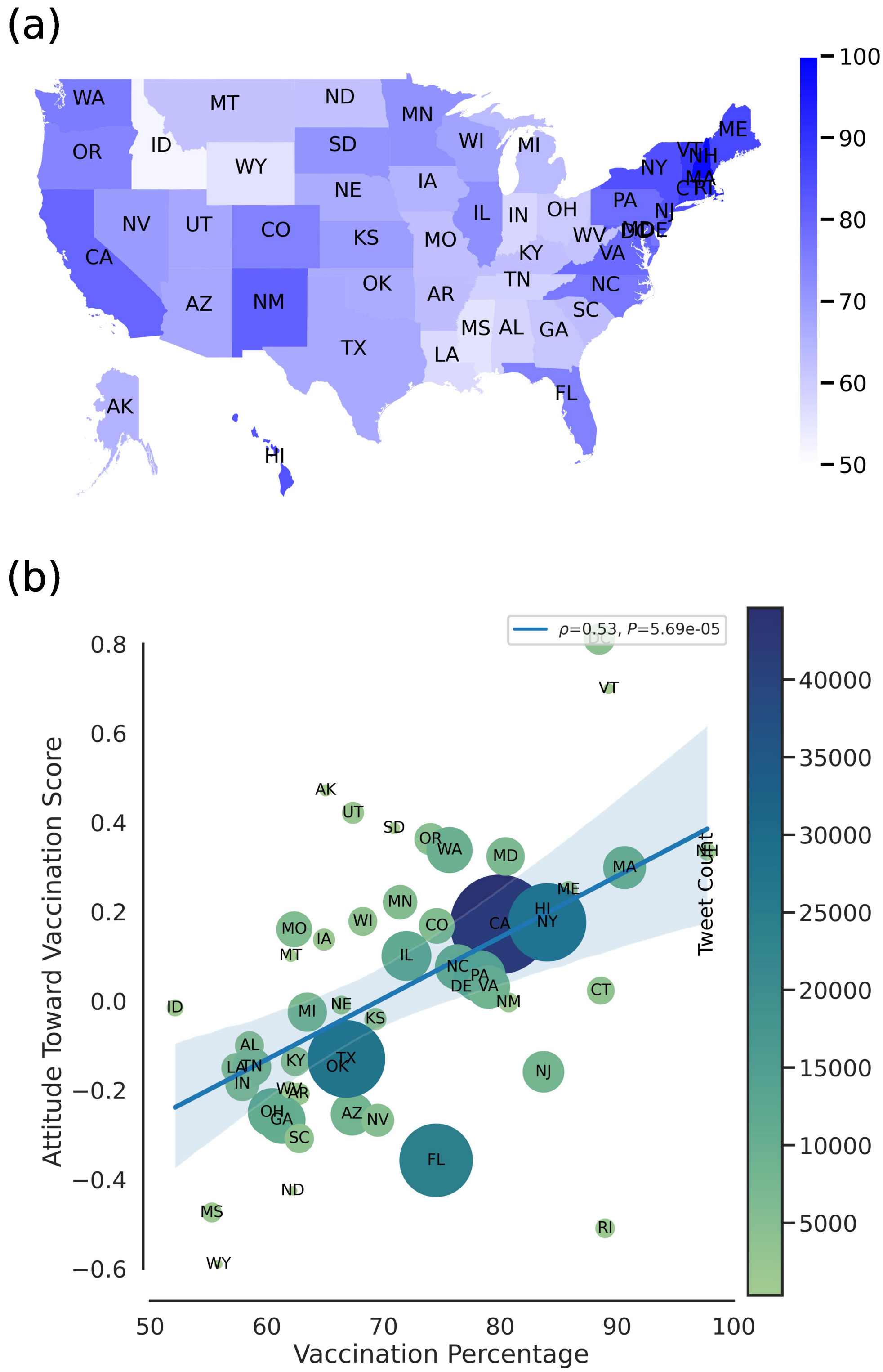

Figure 4: (a) The map displays the vaccination percentage of US states as of December 31st, 2021. (b) This scatter plot presents the correlation between the ATV score of each state and the percentage of eligible population vaccinated in the state. A high Spearman's correlation coefficient indicates that attitude on social media is predictive of the actual vaccination rate. The size and color of the points represent the tweet count of each state.

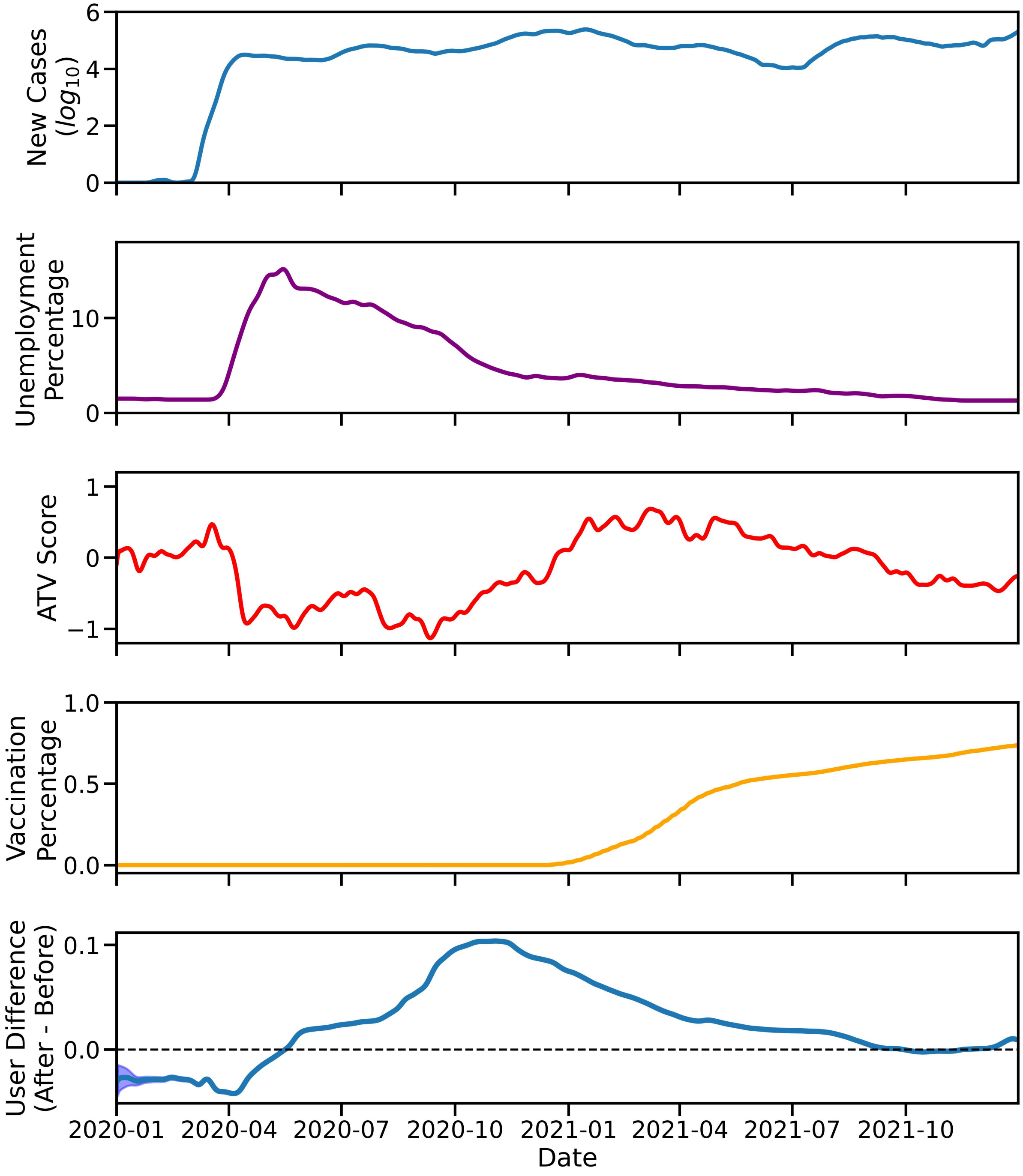

Figure 5: This figure displays the trend of the ATV score and user attitude difference, along with other relevant plots such as COVID data (New Cases and Vaccination Percentage Timeline) and Socio-Economic Parameters (Unemployment Percentage).

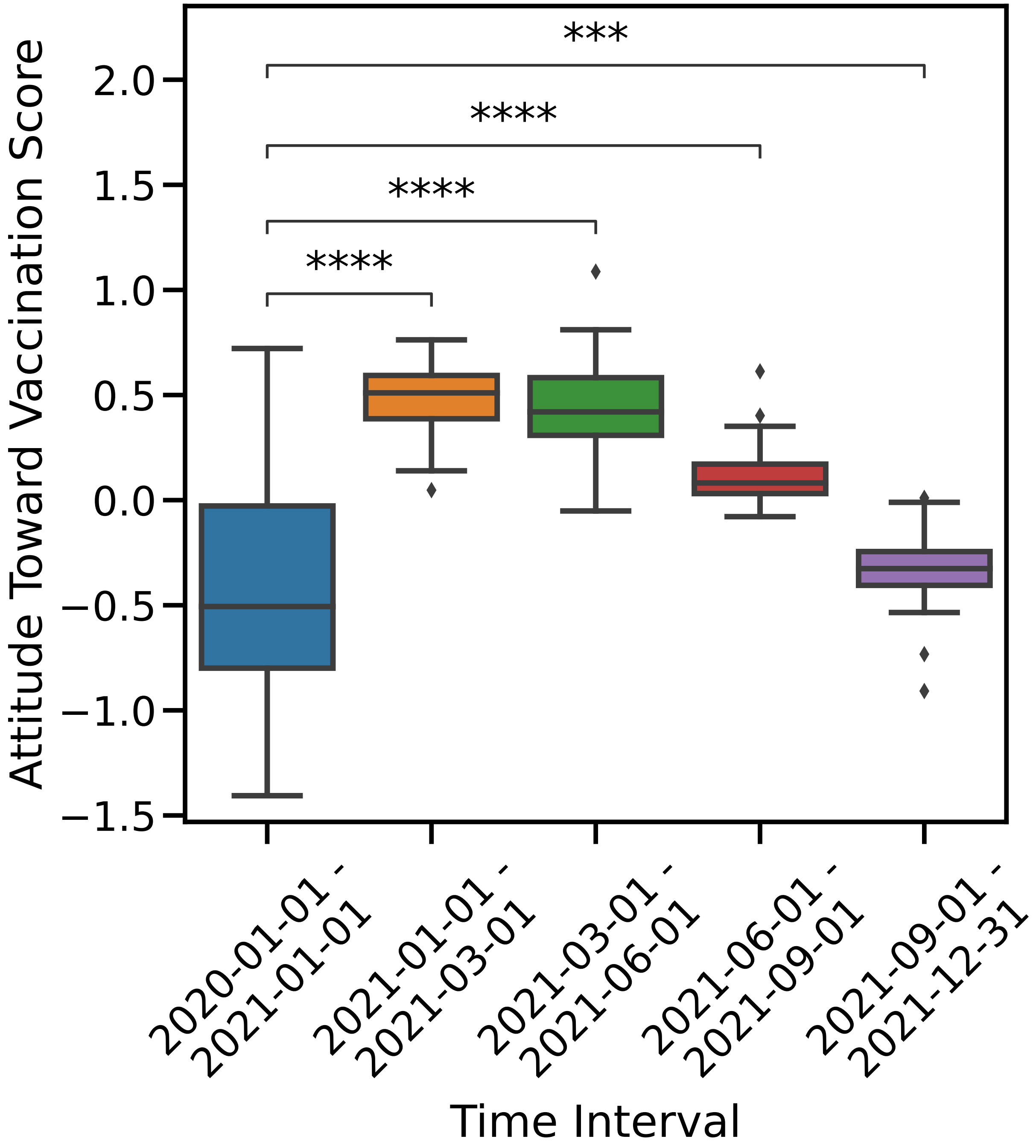

Figure 6: The figure depicts how the Attitude Towards Vaccination (ATV) score changes over time at three-month intervals. To accommodate the lack of tweets in 2020, we plotted the ATV scores for that year in a single box plot until 2021. The Mann-Whitney U test was used to calculate the statistical significance of each box plot with Bonferroni correction.

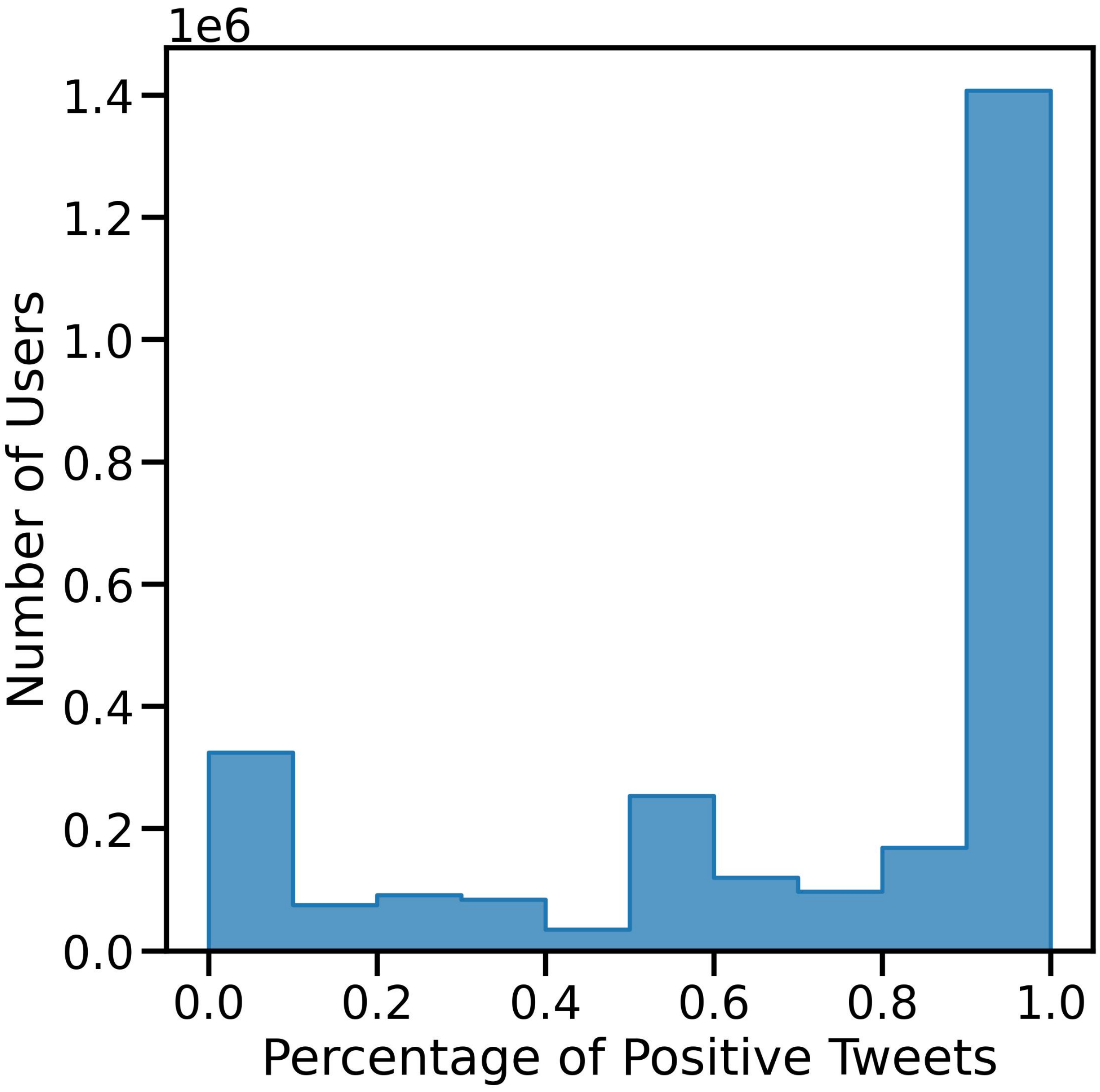

Figure 7: The histogram displays the percentage of positive tweets from users who posted multiple tweets, revealing polarization into two stances.

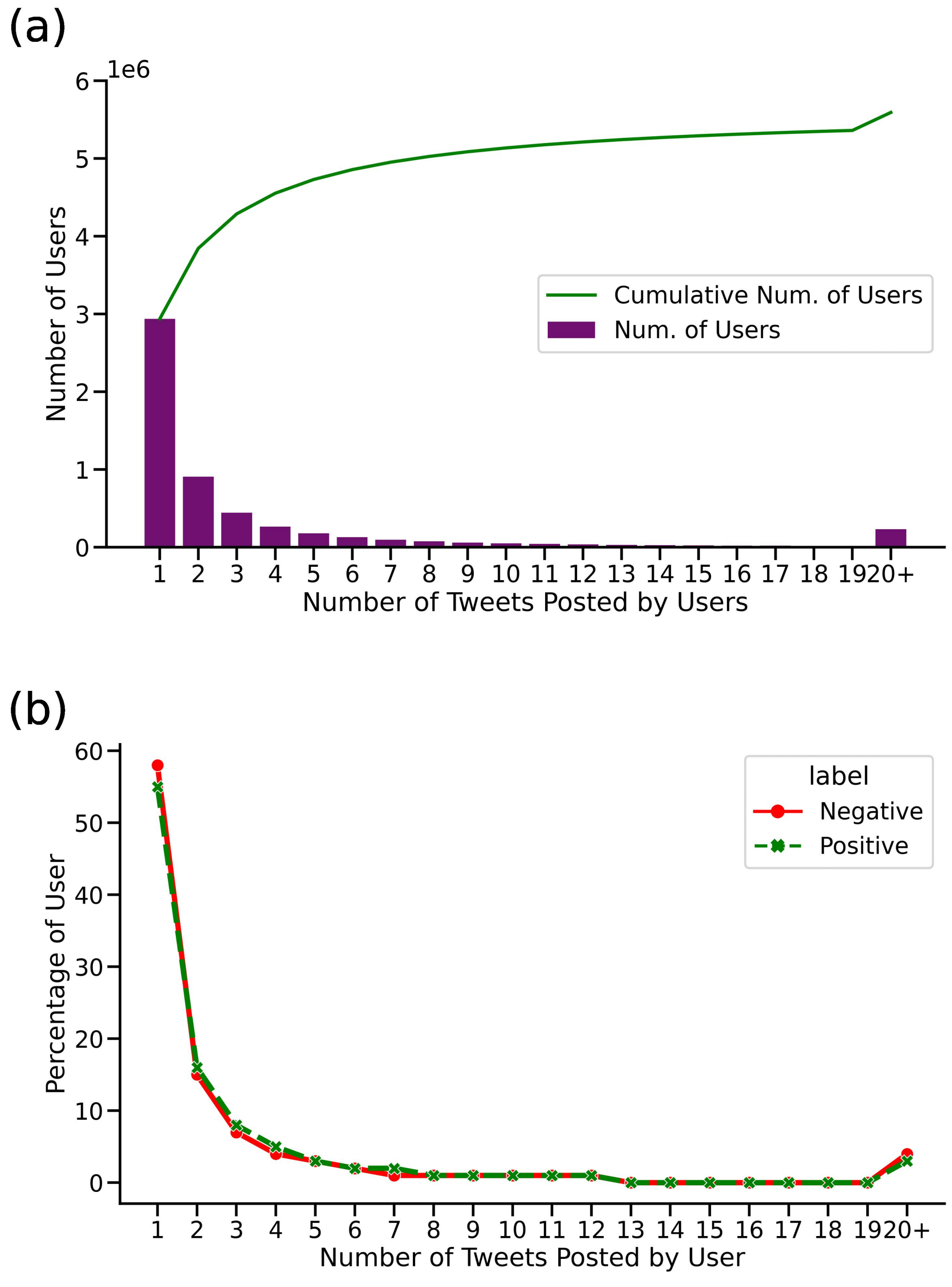

Figure 8: (a) Number of positive or negative tweets sent by users as a histogram and cumulative numbers. Approximately, half of the users sent only one tweet, while the other half sent multiple tweets. (b) The percentage of users based on the number of tweets they posted by tweet attitude. It reveals that users with negative and positive attitudes did not differ in terms of the number of tweets they posted.

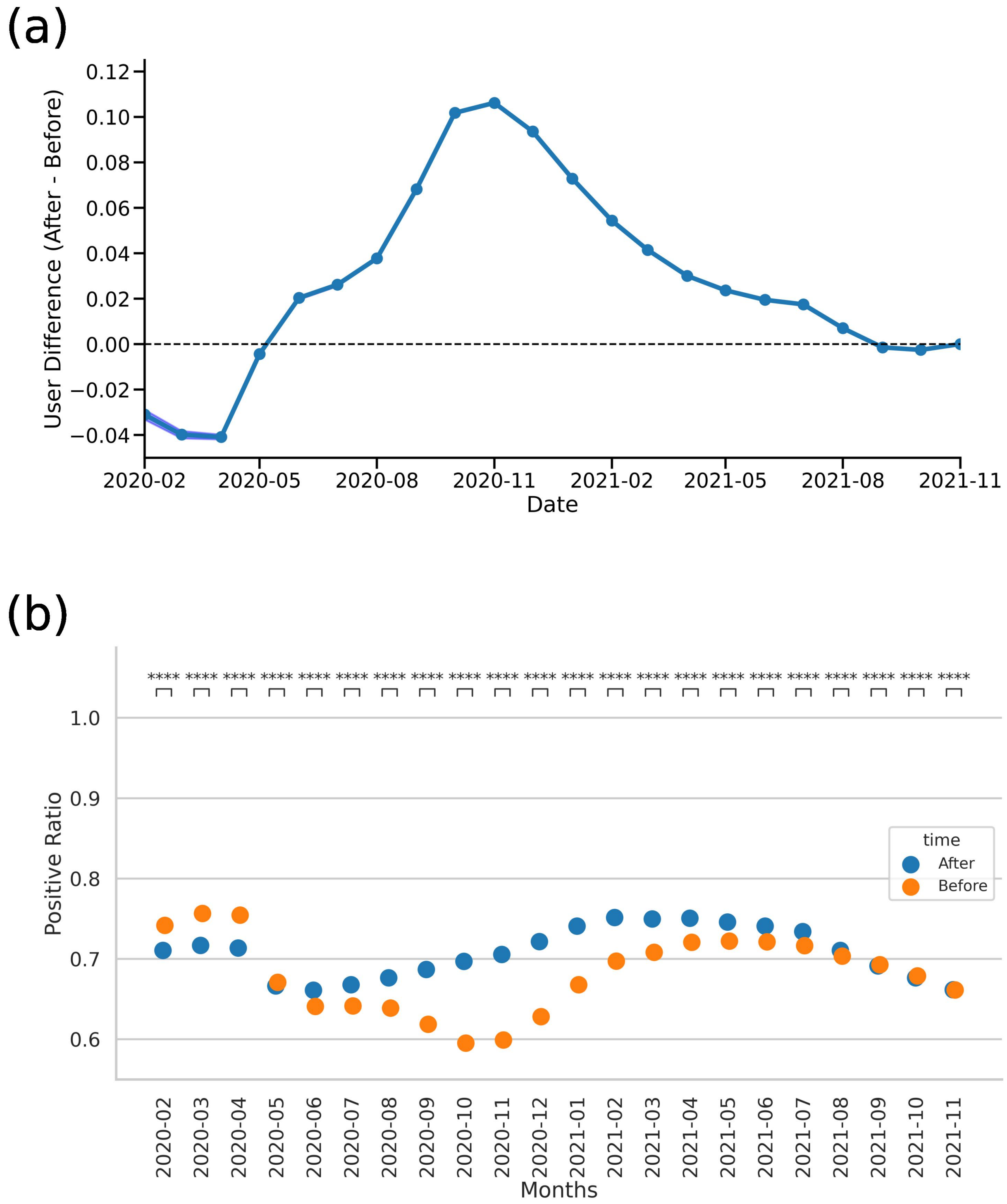

Figure 9: To examine the monthly difference in user attitudes, we selected users who posted at least one tweet before and after a certain time period and calculated their attitude difference and the standard error of the difference. The figure includes two parts: (a) percentage of users' attitude change in each month, with points on the line indicating the change of average attitude towards a more positive (higher values) or negative attitude (lower values) after each month; and (b) the average monthly positive ratio of each user, with changes before and after each month presented. In November 2020, there was a roughly 11% shift towards a positive attitude. We calculated the p-values using the Wilcoxon Test and corrected them using the Bonferroni method.

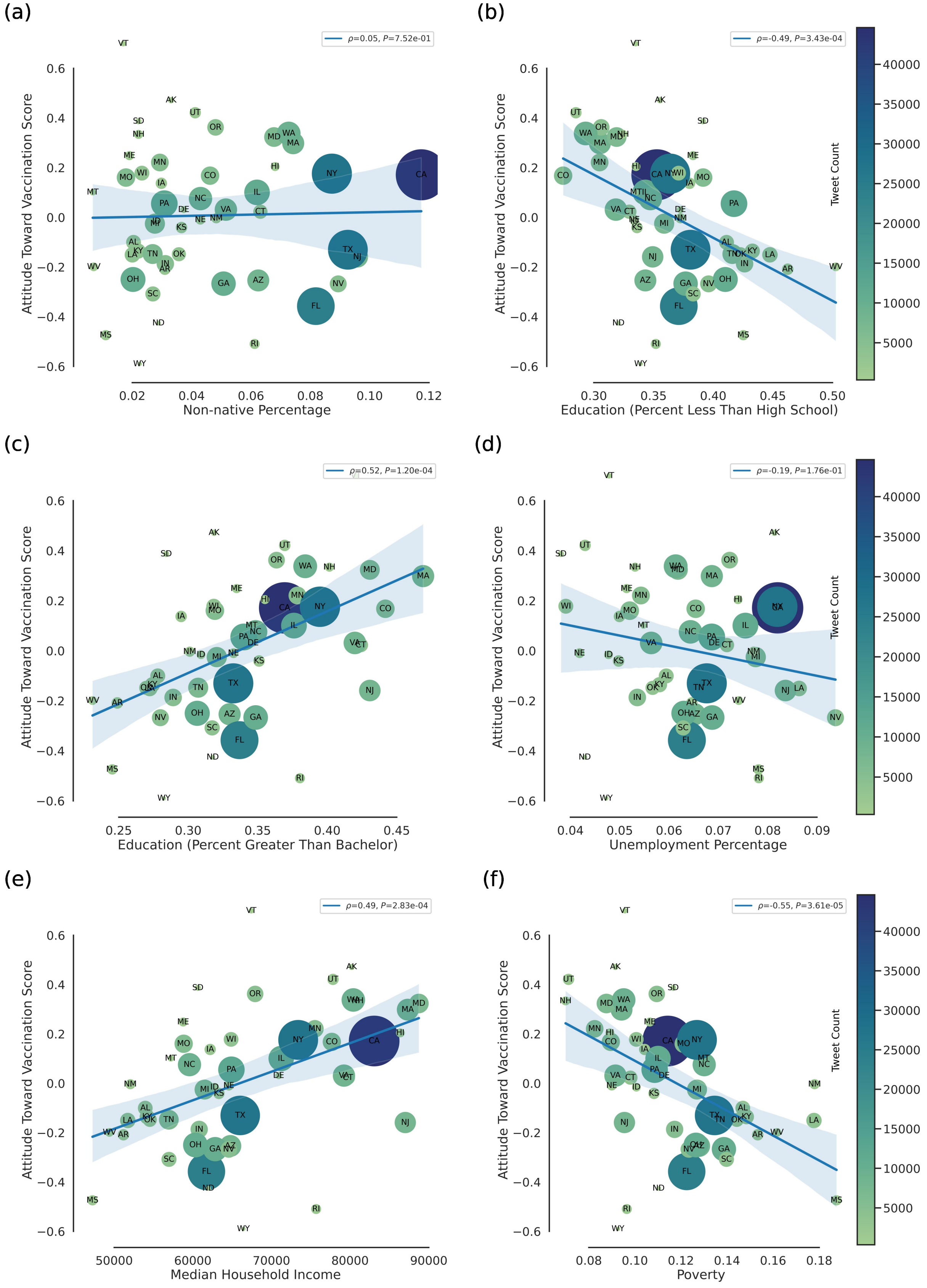
(a)
ρ=0.05, P=7.52e-01
Attitude Toward Vaccination Score
Non-native Percentage
(b)
ρ=-0.49, P=3.43e-04
Attitude Toward Vaccination Score
Education (Percent Less Than High School)
Tweet Count
(c)
ρ=0.52, P=1.20e-04
Attitude Toward Vaccination Score
Education (Percent Greater Than Bachelor)
(d)
ρ=-0.19, P=1.76e-01
Attitude Toward Vaccination Score
Unemployment Percentage
Tweet Count
(e)
ρ=0.49, P=2.83e-04
Attitude Toward Vaccination Score
Median Household Income
(f)
ρ=-0.55, P=3.61e-05
Attitude Toward Vaccination Score
Poverty
Tweet Count

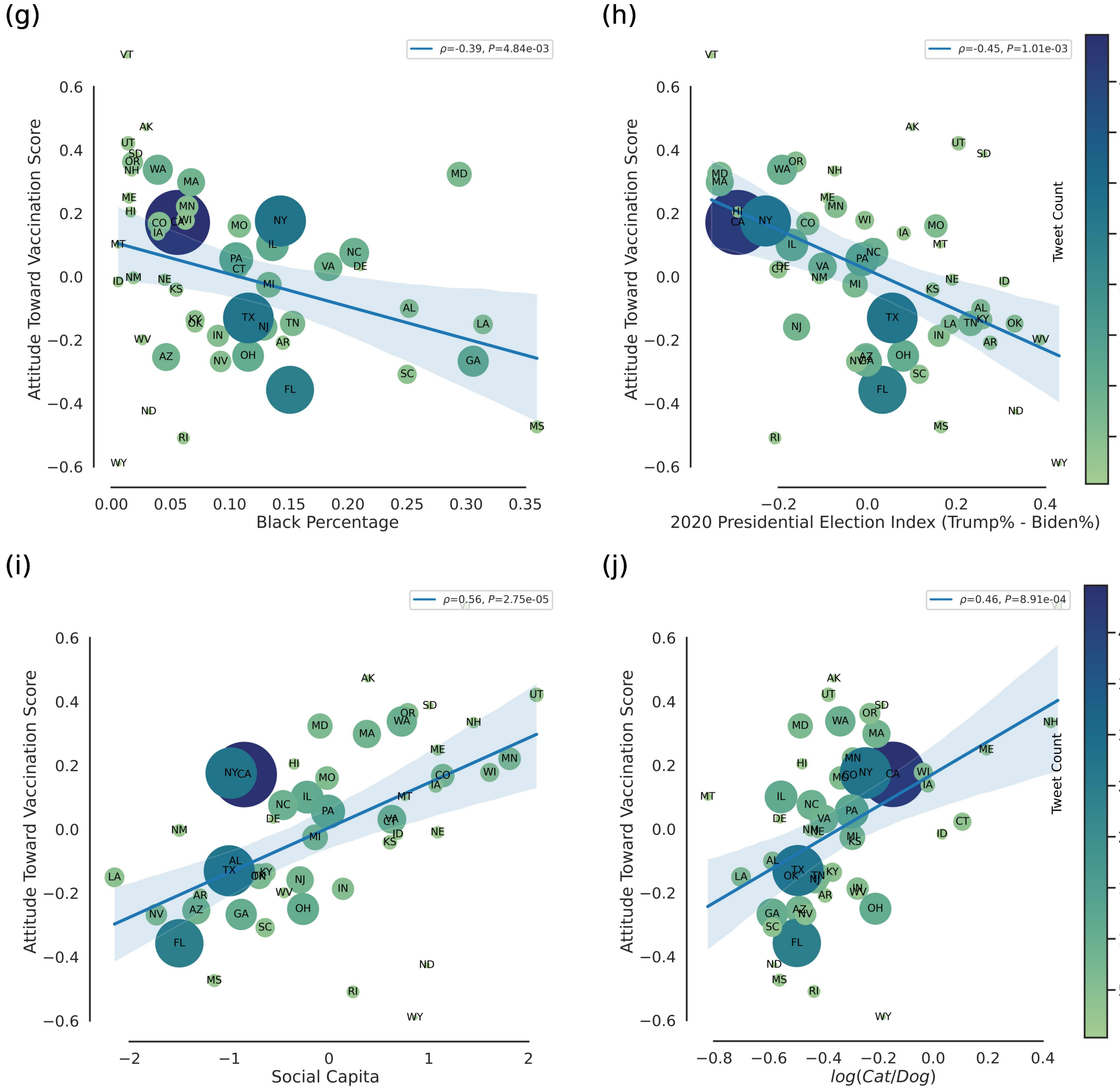

Figure 10: (a-j) Correlation between ATV score and Socio-Economic parameters.

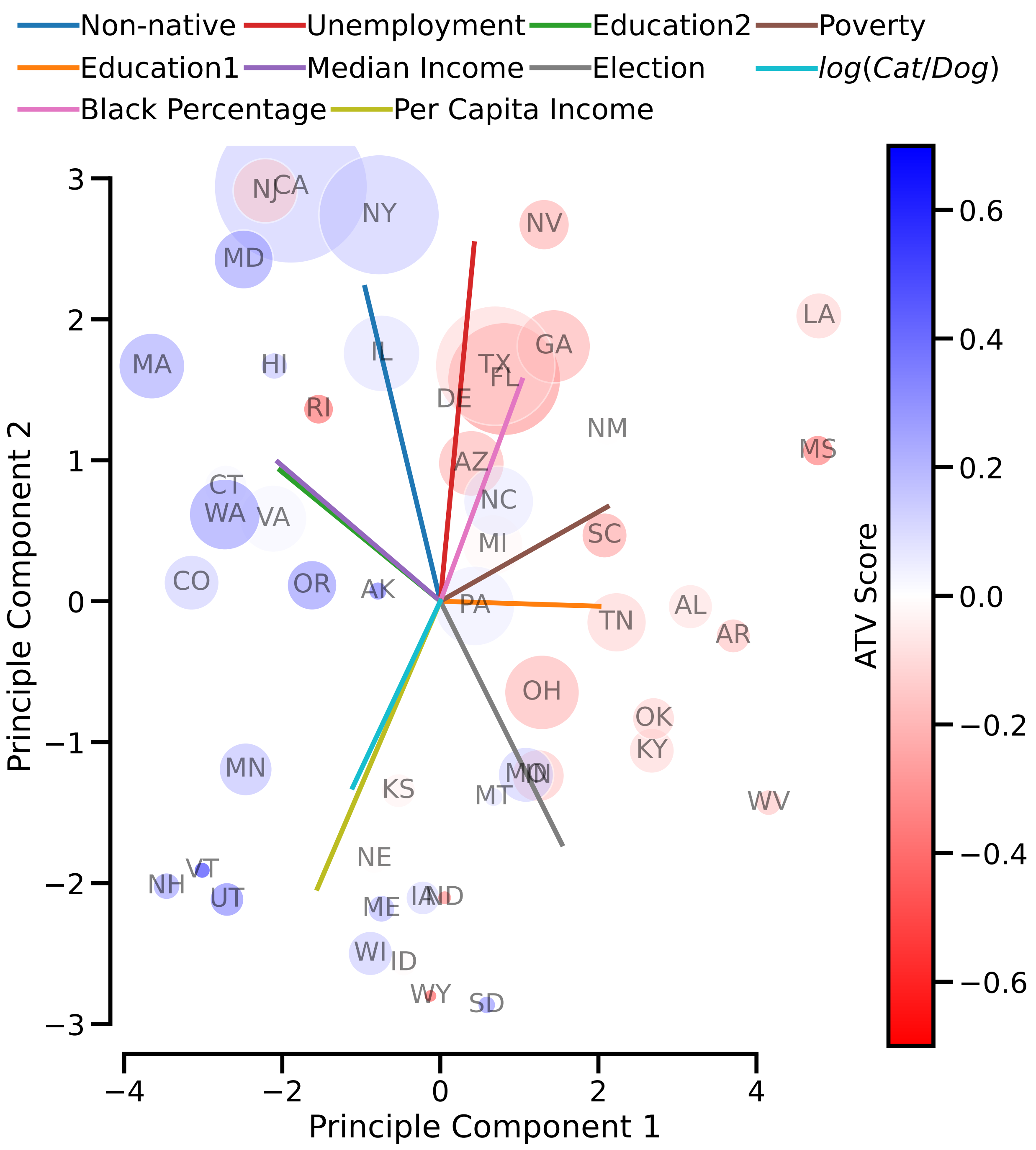

Figure 11: Bi-plot displaying the first two principal components of the socio-economic of the states, where the direction of each socio-economic is indicated with a line.

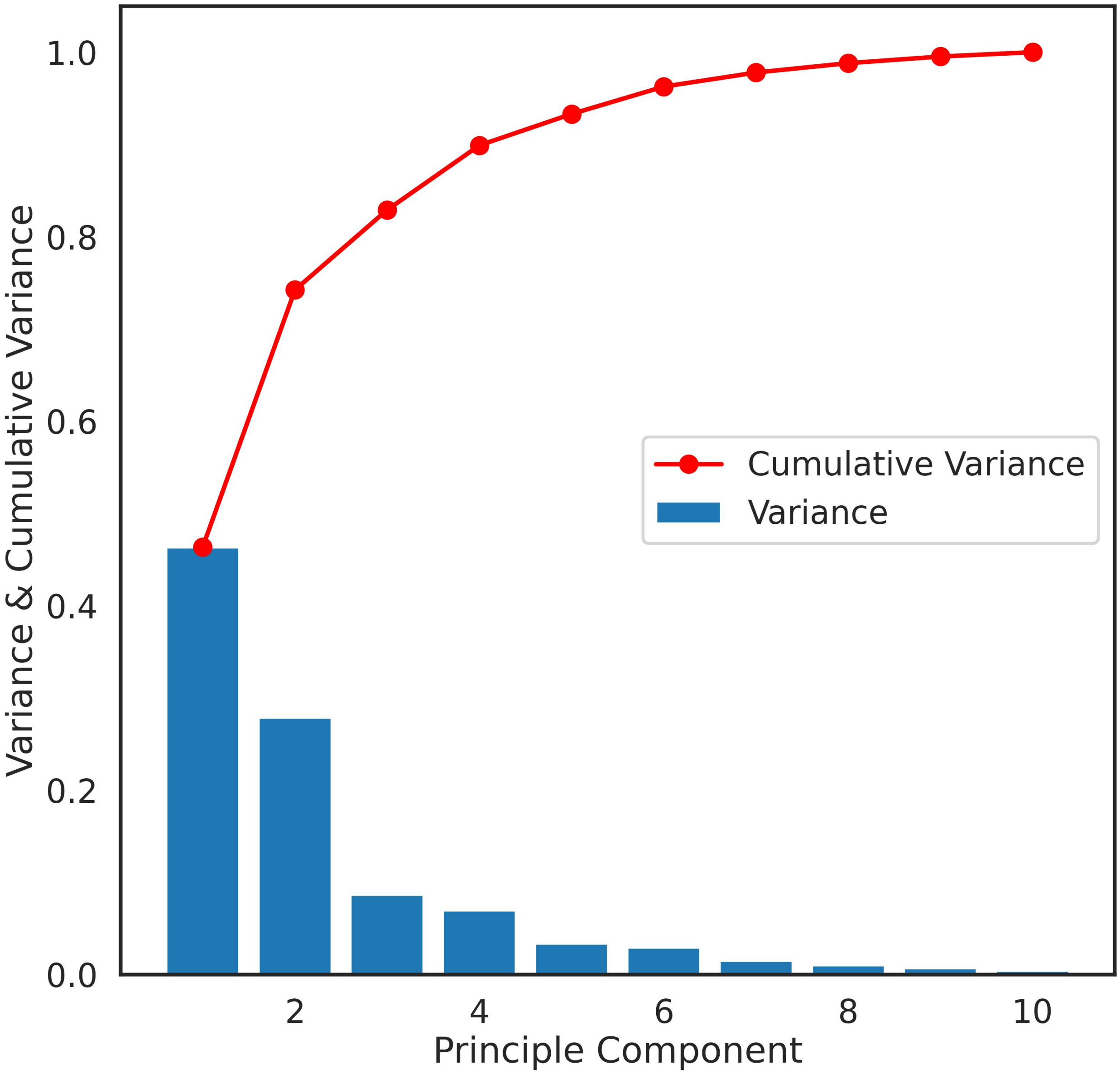

Figure 12: Percentage of variance explained by each principal component displayed as a histogram and cumulative plot. Over 75% of the variance is explained by the top two principal components.

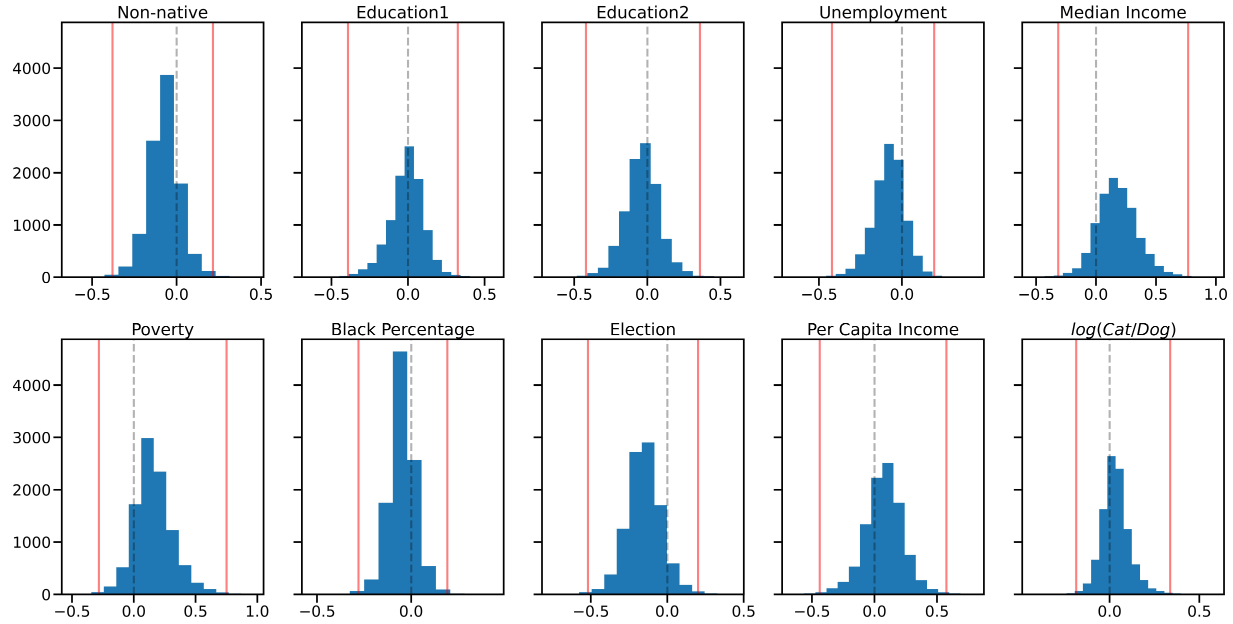

Figure 13: The distribution of coefficients for each socio-economic parameter based on bootstrapped analysis of Partial Least Squares Regression (PLSRegression). The figure includes a dashed line indicating the null hypothesis that the coefficient is zero, and red lines representing the confidence intervals for the coefficient. The null hypothesis (zero coefficient) is contained within these intervals.

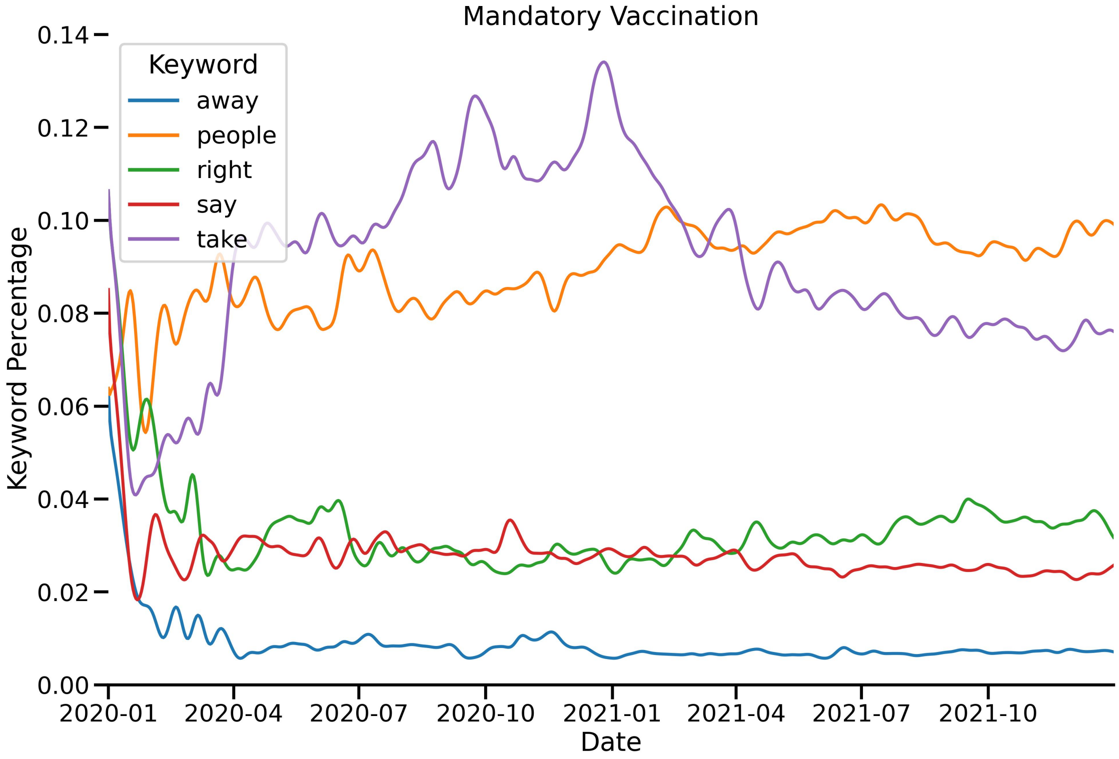
Mandatory Vaccination
Keyword
away
people
right
say
take
Keyword Percentage
0.14
0.12
0.10
0.08
0.06
0.04
0.02
0.00
2020-01
2020-04
2020-07
2020-10
2021-01
2021-04
2021-07
2021-10
Date

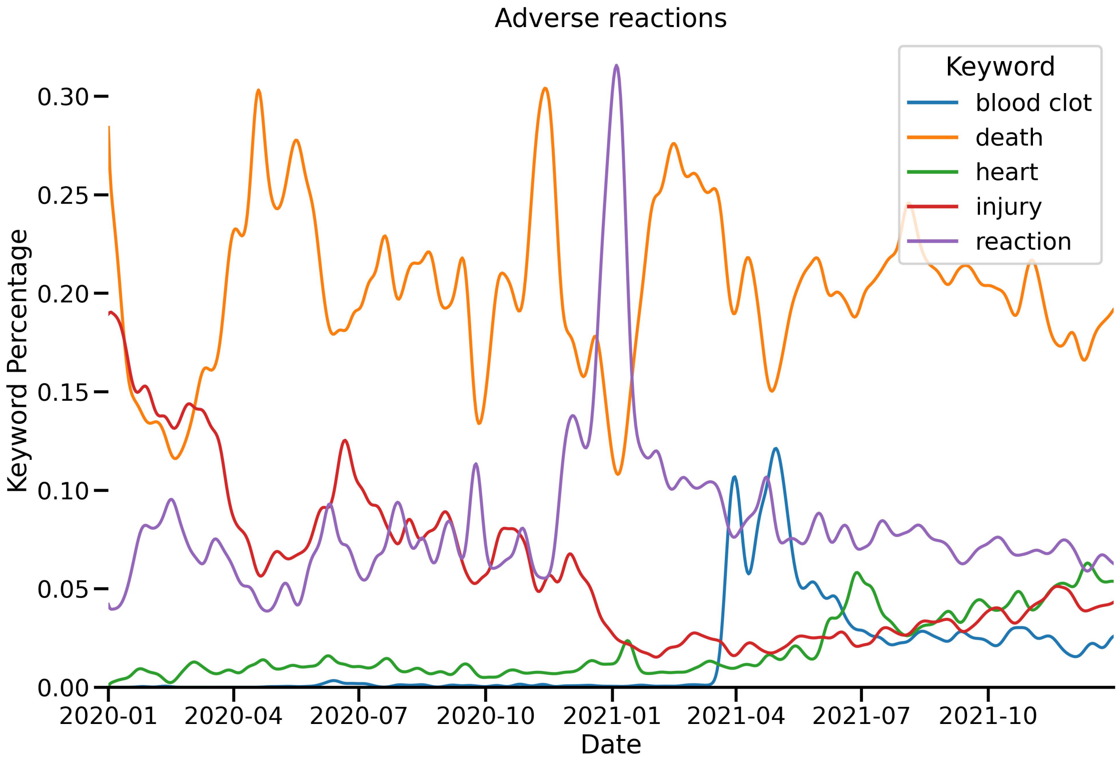
Adverse reactions
Keyword
blood clot
death
heart
injury
reaction
Keyword Percentage
0.30
0.25
0.20
0.15
0.10
0.05
0.00
2020-01
2020-04
2020-07
2020-10
2021-01
2021-04
2021-07
2021-10
Date

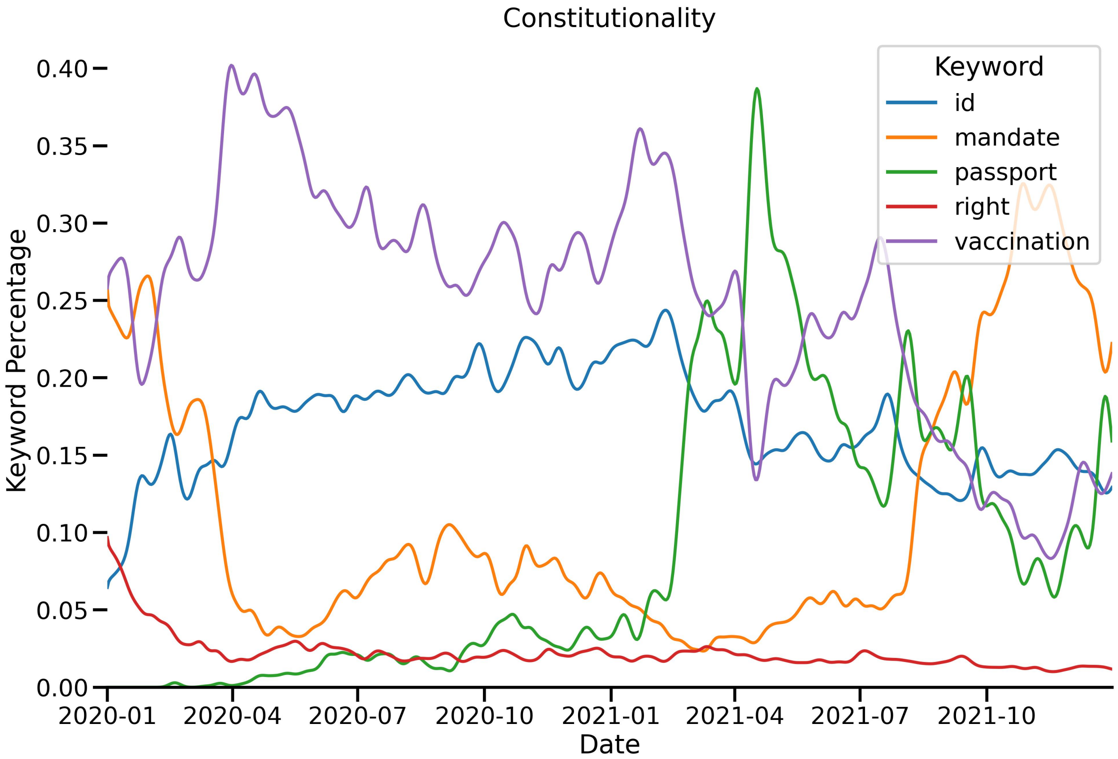
Constitutionality
Keyword
id
mandate
passport
right
vaccination
Keyword Percentage
0.40
0.35
0.30
0.25
0.20
0.15
0.10
0.05
0.00
2020-01
2020-04
2020-07
2020-10
2021-01
2021-04
2021-07
2021-10
Date

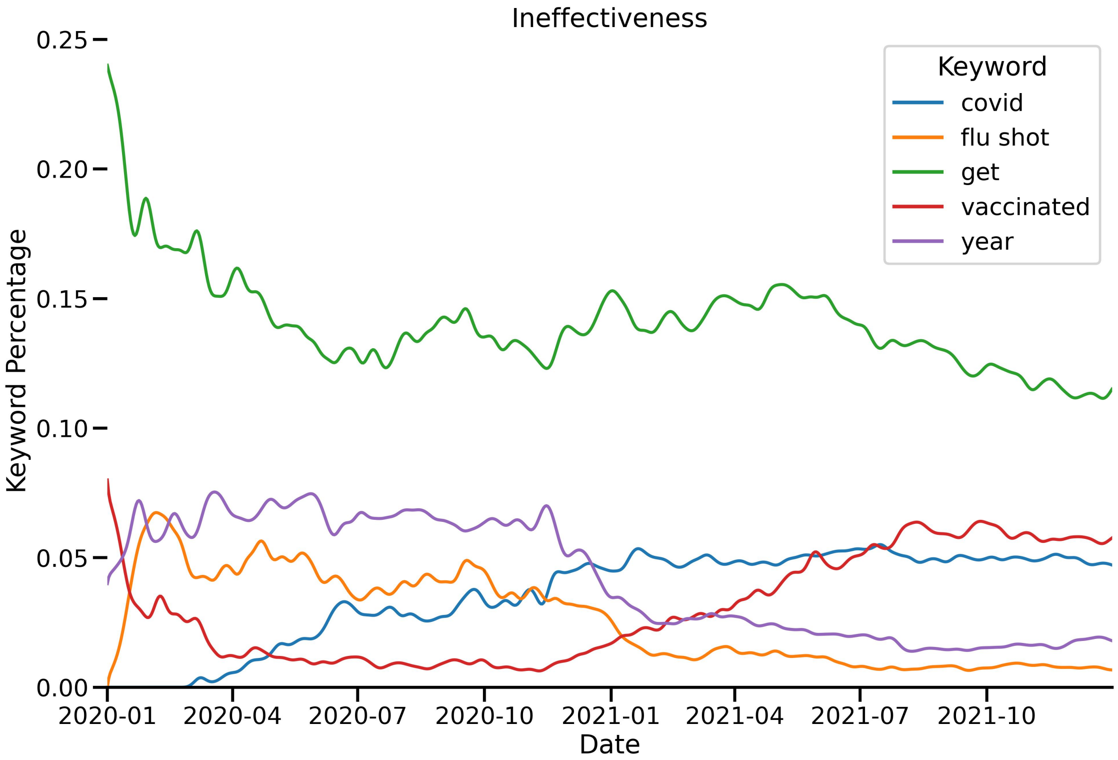
Ineffectiveness
Keyword
covid
flu shot
get
vaccinated
year
Keyword Percentage
0.25
0.20
0.15
0.10
0.05
0.00
2020-01
2020-04
2020-07
2020-10
2021-01
2021-04
2021-07
2021-10
Date

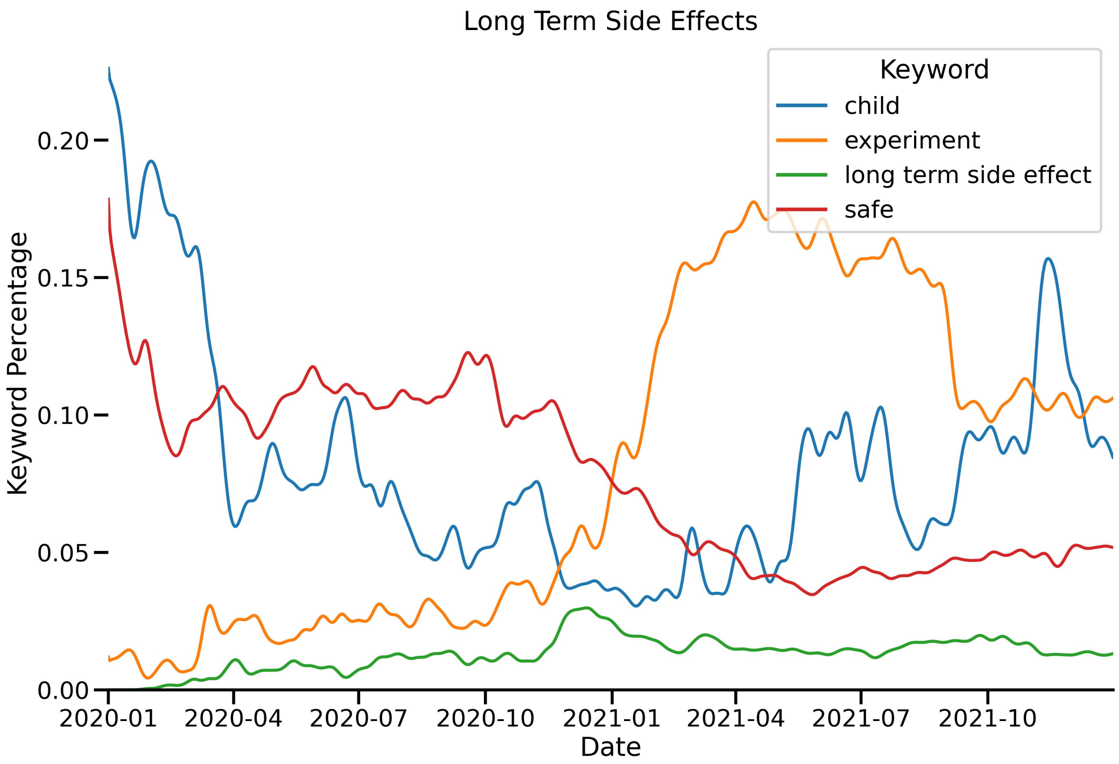
Long Term Side Effects
Keyword
child
experiment
long term side effect
safe
Keyword Percentage
0.00
0.05
0.10
0.15
0.20
2020-01
2020-04
2020-07
2020-10
2021-01
2021-04
2021-07
2021-10
Date

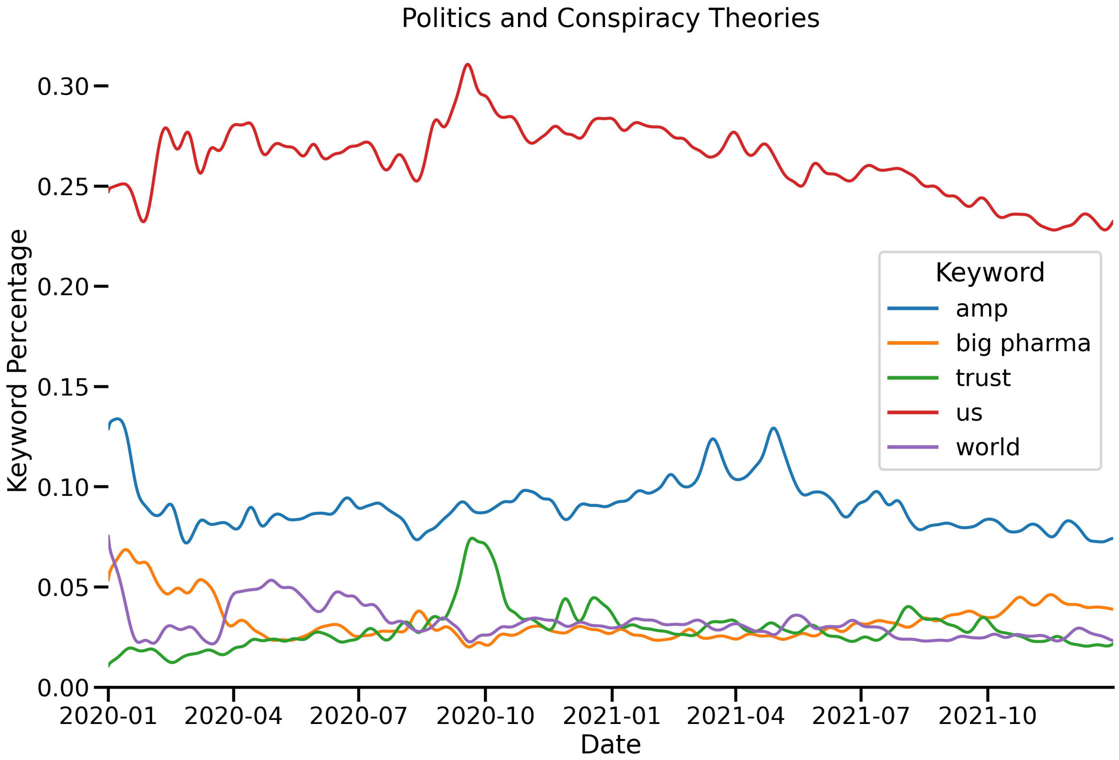
Politics and Conspiracy Theories
Keyword
amp
big pharma
trust
us
world
Keyword Percentage
0.00
0.05
0.10
0.15
0.20
0.25
0.30
2020-01
2020-04
2020-07
2020-10
2021-01
2021-04
2021-07
2021-10
Date

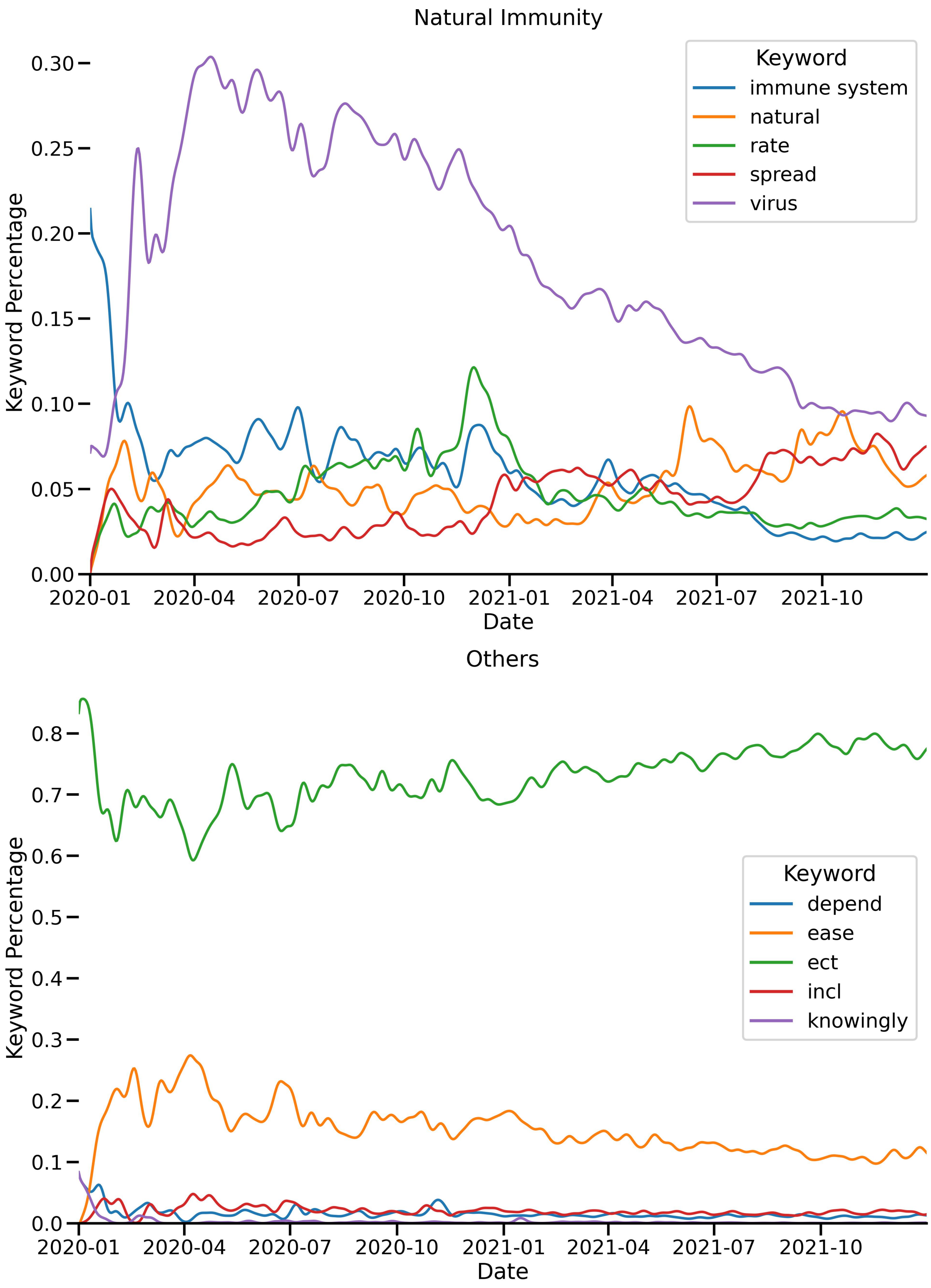

Figure 14: The breakdown of tweets by themes.

Table 1:

| name | code | count | ratio |
|---|---|---|---|
| United States | US | 618752 | 0.486684359 |
| United Kingdom | GB | 197172 | 0.155087221 |
| India | IN | 129941 | 0.102206138 |
| Canada | CA | 84584 | 0.066530225 |
| Australia | AU | 41550 | 0.032681486 |
| South Africa | ZA | 28090 | 0.022094415 |
| Ireland | IE | 22299 | 0.017539458 |
| Philippines | PH | 11243 | 0.008843272 |
| Malaysia | MY | 10617 | 0.008350887 |
| Nigeria | NG | 8429 | 0.006629898 |
| New Zealand | NZ | 7200 | 0.005663218 |
| Pakistan | PK | 7128 | 0.005606586 |
| Germany | DE | 5478 | 0.004308765 |
| Kenya | KE | 5423 | 0.004265504 |
| Netherlands | NL | 4394 | 0.003456136 |
| Spain | ES | 4120 | 0.003240619 |
| Jamaica | JM | 3909 | 0.003074655 |
| France | FR | 3693 | 0.002904759 |
| Italy | IT | 3269 | 0.002571258 |
| United Arab Emirates | AE | 3165 | 0.002489456 |
| Uganda | UG | 2994 | 0.002354955 |
| Thailand | TH | 2934 | 0.002307761 |
| Ghana | GH | 2872 | 0.002258995 |
| Saudi Arabia | SA | 2357 | 0.001853917 |
| Japan | JP | 2313 | 0.001819309 |
| Mexico | MX | 2312 | 0.001818522 |
| Belgium | BE | 2307 | 0.001814589 |
| Singapore | SG | 1951 | 0.001534575 |
| Brazil | BR | 1795 | 0.001411872 |
| Indonesia | "ID" | 1788 | 0.001406366 |
| Trinidad and Tobago | TT | 1767 | 0.001389848 |
| Sri Lanka | LK | 1625 | 0.001278157 |
| Switzerland | CH | 1625 | 0.001278157 |
| Botswana | BW | 1573 | 0.001237256 |
| Sweden | SE | 1484 | 0.001167252 |
| Israel | IL | 1467 | 0.001153881 |
| China | CN | 1192 | 0.000937577 |
| Zimbabwe | ZW | 1170 | 0.000920273 |
| Nepal | NP | 1028 | 0.000808582 |
| Denmark | DK | 996 | 0.000783412 |
| Poland | PL | 955 | 0.000751163 |
| Portugal | PT | 925 | 0.000727566 |
| Lebanon | LB | 923 | 0.000725993 |
| Bangladesh | BD | 915 | 0.000719701 |
| Norway | NO | 874 | 0.000687452 |
| Hong Kong | HK | 857 | 0.00067408 |
| Zambia | ZM | 857 | 0.00067408 |
| Maldives | MV | 848 | 0.000667001 |
| Greece | GR | 793 | 0.000623741 |
| Korea, Republic of | KR | 788 | 0.000619808 |
| Turkey | TR | 723 | 0.000568681 |
| Colombia | CO | 719 | 0.000565535 |
| Taiwan, Province of China | TW | 716 | 0.000563176 |
| Austria | AT | 712 | 0.000560029 |
| Russian Federation | RU | 638 | 0.000501824 |
| Bahamas | BS | 635 | 0.000499464 |
| Viet Nam | VN | 616 | 0.00048452 |
| Cambodia | KH | 610 | 0.0004798 |
| Fiji | FJ | 603 | 0.000474294 |
| Finland | FI | 587 | 0.00046171 |

| name | code | count | ratio |
|---|---|---|---|
| Qatar | QA | 572 | 0.000449911 |
| Barbados | BB | 561 | 0.000441259 |
| Cyprus | CY | 514 | 0.000404291 |
| Argentina | AR | 509 | 0.000400358 |
| Tanzania, United Republic of | TZ | 497 | 0.000390919 |
| Malawi | MW | 461 | 0.000362603 |
| Cuba | CU | 437 | 0.000343726 |
| Egypt | EG | 411 | 0.000323275 |
| Kuwait | KW | 398 | 0.00031305 |
| Antigua and Barbuda | AG | 396 | 0.000311477 |
| Romania | RO | 388 | 0.000305185 |
| Bahrain | BH | 371 | 0.000291813 |
| Czech Republic | CZ | 365 | 0.000287094 |
| Panama | PA | 359 | 0.000282374 |
| Serbia | RS | 329 | 0.000258778 |
| Hungary | HU | 312 | 0.000245406 |
| Costa Rica | CR | 310 | 0.000243833 |
| Rwanda | RW | 289 | 0.000227315 |
| Chile | CL | 282 | 0.000221809 |
| Oman | OM | 264 | 0.000207651 |
| Ethiopia | ET | 246 | 0.000193493 |
| Saint Lucia | LC | 239 | 0.000187987 |
| Bermuda | BM | 238 | 0.000187201 |
| Dominican Republic | DO | 235 | 0.000184841 |
| Croatia | HR | 232 | 0.000182481 |
| Isle of Man | IM | 227 | 0.000178549 |
| Mauritius | MU | 226 | 0.000177762 |
| Papua New Guinea | PG | 219 | 0.000172256 |
| Ecuador | EC | 212 | 0.00016675 |
| Lesotho | LS | 209 | 0.000164391 |
| Malta | MT | 208 | 0.000163604 |
| Peru | PE | 207 | 0.000162818 |
| Gibraltar | GI | 201 | 0.000158098 |
| Brunei Darussalam | BN | 192 | 0.000151019 |
| Iraq | IQ | 184 | 0.000144727 |
| Ukraine | UA | 179 | 0.000140794 |
| Gambia | GM | 176 | 0.000138434 |
| Anguilla | AI | 173 | 0.000136075 |
| Georgia | GE | 173 | 0.000136075 |
| Somalia | SO | 172 | 0.000135288 |
| Venezuela, Bolivarian Republic of | VE | 165 | 0.000129782 |
| Jordan | JO | 162 | 0.000127422 |
| Kazakhstan | KZ | 162 | 0.000127422 |
| Iran, Islamic Republic of | IR | 161 | 0.000126636 |
| Myanmar | MM | 154 | 0.00012113 |
| Swaziland | SZ | 150 | 0.000117984 |
| Bulgaria | BG | 147 | 0.000115624 |
| Iceland | IS | 146 | 0.000114837 |
| Saint Vincent and the Grenadines | VC | 140 | 0.000110118 |
| Albania | AL | 139 | 0.000109332 |
| Cayman Islands | KY | 138 | 0.000108545 |
| Uruguay | UY | 134 | 0.000105399 |
| Yemen | YE | 132 | 0.000103826 |
| Luxembourg | LU | 130 | 0.000102253 |
| Honduras | HN | 129 | 0.000101466 |
| Lithuania | LT | 126 | 9.91063E-05 |
| Morocco | MA | 124 | 9.75332E-05 |
| Latvia | LV | 121 | 9.51735E-05 |
| Tunisia | TN | 121 | 9.51735E-05 |
| Slovenia | SI | 119 | 9.36004E-05 |

| name | code | count | ratio |
|---|---|---|---|
| Estonia | EE | 115 | 9.04542E-05 |
| Guatemala | GT | 114 | 8.96676E-05 |
| Liberia | LR | 96 | 7.55096E-05 |
| Sierra Leone | SL | 95 | 7.4723E-05 |
| Grenada | GD | 90 | 7.07902E-05 |
| Cameroon | CM | 86 | 6.7644E-05 |
| El Salvador | SV | 82 | 6.44978E-05 |
| Guyana | GY | 79 | 6.21381E-05 |
| Slovakia | SK | 79 | 6.21381E-05 |
| Aruba | AW | 76 | 5.97784E-05 |
| Belize | BZ | 72 | 5.66322E-05 |
| Mongolia | MN | 71 | 5.58456E-05 |
| Mozambique | MZ | 71 | 5.58456E-05 |
| Afghanistan | AF | 70 | 5.50591E-05 |
| Senegal | SN | 70 | 5.50591E-05 |
| Bhutan | BT | 69 | 5.42725E-05 |
| Congo, the Democratic Republic | CD | 65 | 5.11263E-05 |
| Macedonia, the Former Y.R. | MK | 65 | 5.11263E-05 |
| Virgin Islands, U.S. | VI | 64 | 5.03397E-05 |
| Turks and Caicos Islands | TC | 63 | 4.95532E-05 |
| Nicaragua | NI | 61 | 4.798E-05 |
| Paraguay | PY | 56 | 4.40473E-05 |
| Bolivia, Plurinational State of | BO | 54 | 4.24741E-05 |
| Guam | GU | 54 | 4.24741E-05 |
| Libya | LY | 53 | 4.16876E-05 |
| Armenia | AM | 49 | 3.85413E-05 |
| Timor-Leste | TL | 49 | 3.85413E-05 |
| Azerbaijan | AZ | 45 | 3.53951E-05 |
| Côte d'Ivoire | CI | 45 | 3.53951E-05 |
| Seychelles | SC | 44 | 3.46086E-05 |
| Montenegro | ME | 43 | 3.3822E-05 |
| Saint Kitts and Nevis | KN | 39 | 3.06758E-05 |
| Algeria | DZ | 38 | 2.98892E-05 |
| Bosnia and Herzegovina | BA | 38 | 2.98892E-05 |
| Moldova, Republic of | MD | 33 | 2.59564E-05 |
| Congo | CG | 31 | 2.43833E-05 |
| Dominica | DM | 31 | 2.43833E-05 |
| Virgin Islands, British | VG | 29 | 2.28102E-05 |
| Haiti | HT | 28 | 2.20236E-05 |
| Northern Mariana Islands | MP | 28 | 2.20236E-05 |
| Angola | AO | 27 | 2.12371E-05 |
| Samoa | WS | 27 | 2.12371E-05 |
| San Marino | SM | 27 | 2.12371E-05 |
| Suriname | SR | 24 | 1.88774E-05 |
| Vanuatu | VU | 24 | 1.88774E-05 |
| Syrian Arab Republic | SY | 23 | 1.80908E-05 |
| Monaco | MC | 22 | 1.73043E-05 |
| Macao | MO | 19 | 1.49446E-05 |
| Kyrgyzstan | KG | 18 | 1.4158E-05 |
| United States M.O.I | UM | 18 | 1.4158E-05 |
| Benin | BJ | 17 | 1.33715E-05 |
| Puerto Rico | PR | 17 | 1.33715E-05 |
| Andorra | AD | 16 | 1.25849E-05 |
| Liechtenstein | LI | 16 | 1.25849E-05 |
| Solomon Islands | SB | 15 | 1.17984E-05 |
| Uzbekistan | UZ | 15 | 1.17984E-05 |
| Lao People's Democratic Republic | LA | 14 | 1.10118E-05 |
| Burkina Faso | BF | 13 | 1.02253E-05 |
| Faroe Islands | FO | 13 | 1.02253E-05 |
| Palau | PW | 13 | 1.02253E-05 |
| Tonga | TO | 13 | 1.02253E-05 |
| Cape Verde | CV | 12 | 9.4387E-06 |
| French Polynesia | PF | 12 | 9.4387E-06 |
| Sint Maarten (Dutch part) | SX | 12 | 9.4387E-06 |

| name | code | count | ratio |
|---|---|---|---|
| Tajikistan | TJ | 12 | 9.4387E-06 |
| Jersey | JE | 11 | 8.65214E-06 |
| Gabon | GA | 10 | 7.86558E-06 |
| Guernsey | GG | 10 | 7.86558E-06 |
| Montserrat | MS | 10 | 7.86558E-06 |
| Burundi | BI | 9 | 7.07902E-06 |
| Central African Republic | CF | 9 | 7.07902E-06 |
| Curaçao | CW | 9 | 7.07902E-06 |
| Saint Martin (French part) | MF | 9 | 7.07902E-06 |
| Togo | TG | 9 | 7.07902E-06 |
| Djibouti | DJ | 8 | 6.29246E-06 |
| Guinea | GN | 8 | 6.29246E-06 |
| Martinique | MQ | 8 | 6.29246E-06 |
| Bonaire, Sint Eustatius and Saba | BQ | 6 | 4.71935E-06 |
| Holy See (Vatican City State) | VA | 6 | 4.71935E-06 |
| Madagascar | MG | 6 | 4.71935E-06 |
| Chad | TD | 5 | 3.93279E-06 |
| Guadeloupe | GP | 5 | 3.93279E-06 |
| Marshall Islands | MH | 5 | 3.93279E-06 |
| Niue | NU | 5 | 3.93279E-06 |
| Sudan | SD | 5 | 3.93279E-06 |
| Belarus | BY | 4 | 3.14623E-06 |
| Equatorial Guinea | GQ | 4 | 3.14623E-06 |
| Greenland | GL | 4 | 3.14623E-06 |
| Korea, Democratic People's Republic | KP | 4 | 3.14623E-06 |
| Saint Barthélemy | BL | 4 | 3.14623E-06 |
| Comoros | KM | 3 | 2.35967E-06 |
| Cook Islands | CK | 3 | 2.35967E-06 |
| Kiribati | KI | 3 | 2.35967E-06 |
| Saint Helena, Ascension and T. da C. | SH | 3 | 2.35967E-06 |
| American Samoa | AS | 2 | 1.57312E-06 |
| Antarctica | AQ | 2 | 1.57312E-06 |
| Falkland Islands (Malvinas) | FK | 2 | 1.57312E-06 |
| Mali | ML | 2 | 1.57312E-06 |
| Mayotte | YT | 2 | 1.57312E-06 |
| Micronesia, Federated States of | FM | 2 | 1.57312E-06 |
| Nauru | NR | 2 | 1.57312E-06 |
| Niger | NE | 2 | 1.57312E-06 |
| Turkmenistan | TM | 2 | 1.57312E-06 |
| Åland Islands | AX | 1 | 7.86558E-07 |
| Christmas Island | CX | 1 | 7.86558E-07 |
| French Guiana | GF | 1 | 7.86558E-07 |
| Guinea-Bissau | GW | 1 | 7.86558E-07 |
| Mauritania | MR | 1 | 7.86558E-07 |
| New Caledonia | NC | 1 | 7.86558E-07 |
| Réunion | RE | 1 | 7.86558E-07 |
| Bouvet Island | BV | 0 | 0 |
| British Indian Ocean Territory | IO | 0 | 0 |
| Cocos (Keeling) Islands | CC | 0 | 0 |
| Eritrea | ER | 0 | 0 |
| French Southern Territories | TF | 0 | 0 |
| Heard Island and McDonald Islands | HM | 0 | 0 |
| Norfolk Island | NF | 0 | 0 |
| Palestine, State of | PS | 0 | 0 |
| Pitcairn | PN | 0 | 0 |
| Saint Pierre and Miquelon | PM | 0 | 0 |
| Sao Tome and Principe | ST | 0 | 0 |
| South Georgia and the South S.I. | GS | 0 | 0 |
| South Sudan | SS | 0 | 0 |
| Svalbard and Jan Mayen | SJ | 0 | 0 |
| Tokelau | TK | 0 | 0 |
| Tuvalu | TV | 0 | 0 |
| Wallis and Futuna | WF | 0 | 0 |
| Western Sahara | EH | 0 | 0 |

Table 2:

| label | precision | recall |
|---|---|---|
| Neutral | 0.992084433 | 0.65849387 |
| Positive | 0.93989071 | 0.866498741 |
| Negative | 0.857142857 | 0.677419355 |

Table 3:

| | precision | recall | f1-score | support |
|---|---|---|---|---|
| Neutral | 0.97 | 0.85 | 0.91 | 571 |
| Positive | 0.82 | 0.96 | 0.88 | 397 |
| Negative | 0.77 | 0.74 | 0.75 | 62 |
| accuracy | | | 0.89 | 1030 |
| macro avg | 0.85 | 0.85 | 0.85 | 1030 |
| weighted avg | 0.9 | 0.89 | 0.89 | 1030 |

Table 4:

| label_change | counts | ratio |
|---|---|---|
| Positive to Positive | 588485 | 0.649233807 |
| Negative to Negative | 142198 | 0.156876979 |
| Negative to Positive | 94163 | 0.103883367 |
| Positive to Negative | 81584 | 0.090005847 |

Table 5:

| State | Tweet Count | P adj | ATV Score |
|---|---|---|---|
| AK | 657 | 8.90447E-06 | 0.471756217 |
| AL | 3779 | 0.011351485 | -0.10041863 |
| AR | 2246 | 3.29141E-05 | -0.208551681 |
| AZ | 8280 | 1.19799E-22 | -0.253782321 |
| CA | 44623 | 5.33886E-42 | 0.171502944 |
| CO | 5794 | 6.07001E-07 | 0.167997364 |
| CT | 3482 | 0.632883957 | 0.022942864 |
| DC | 4417 | 3.44219E-79 | 0.8105628 |
| DE | 847 | 0.789082031 | 0.031302454 |
| FL | 24336 | 6.76491E-122 | -0.357291199 |
| GA | 10252 | 3.10715E-30 | -0.266782302 |
| HI | 1374 | 0.003259278 | 0.204561713 |
| IA | 2217 | 0.011808379 | 0.137276937 |
| "ID" | 1299 | 0.849750857 | -0.015809699 |
| IL | 11236 | 2.92215E-05 | 0.100684802 |
| IN | 5202 | 1.99568E-08 | -0.186645667 |
| KS | 2225 | 0.462111036 | -0.040858535 |
| KY | 3896 | 0.000424907 | -0.135667586 |
| LA | 4097 | 6.04206E-05 | -0.150655316 |
| MA | 8311 | 1.18295E-25 | 0.29857051 |
| MD | 6665 | 3.76726E-24 | 0.323542583 |
| ME | 1413 | 0.000323389 | 0.248632772 |
| MI | 6951 | 0.428105166 | -0.025223656 |
| MN | 5375 | 3.12453E-10 | 0.220998253 |
| MO | 5839 | 1.72973E-06 | 0.160396999 |
| MS | 1807 | 4.36279E-19 | -0.474139401 |
| MT | 709 | 0.326892517 | 0.101143131 |
| NC | 9370 | 0.003821649 | 0.075708428 |
| ND | 393 | 0.000226868 | -0.425525186 |
| NE | 1624 | 0.899687276 | -0.009502075 |
| NH | 1389 | 2.40411E-06 | 0.334188529 |
| NJ | 7863 | 5.51062E-09 | -0.158854963 |
| NM | 1763 | 0.954566869 | -0.003554962 |
| NV | 4893 | 1.27853E-15 | -0.26755032 |
| NY | 27577 | 3.40531E-29 | 0.176041901 |
| OH | 10677 | 1.13831E-27 | -0.249948957 |
| OK | 3399 | 0.000319267 | -0.148272009 |
| OR | 4722 | 2.18641E-21 | 0.36236741 |
| PA | 12164 | 0.01646002 | 0.055224968 |
| RI | 1734 | 2.87262E-21 | -0.509497181 |
| SC | 3913 | 9.37585E-17 | -0.308218329 |
| SD | 618 | 0.000319267 | 0.387028113 |
| TN | 6786 | 5.50772E-07 | -0.147544189 |
| TX | 27141 | 6.07314E-18 | -0.130145282 |
| UT | 2207 | 9.77122E-14 | 0.421307139 |
| VA | 8623 | 0.280125619 | 0.031337135 |
| VT | 517 | 2.76846E-08 | 0.699645135 |
| WA | 9611 | 1.22354E-36 | 0.3378939 |
| WI | 3769 | 1.98307E-05 | 0.178159092 |
| WV | 1295 | 0.002745823 | -0.199078452 |
| WY | 326 | 2.33857E-06 | -0.589826295 |

Table 6:

| Parameter | PC1 | PC2 |
|---|---|---|
| Non-native Percentage | -0.190643915 | 0.445307532 |
| Education (Percent Less Than High School) | 0.401486314 | -0.007066115 |
| Education (Percent Greater Than Bachelor) | -0.405721225 | 0.185800244 |
| Unemployment Percentage | 0.085779135 | 0.507289331 |
| Median Household Income | -0.41090577 | 0.197402667 |
| Poverty | 0.422411252 | 0.13392345 |
| Black Percentage | 0.206902313 | 0.313861901 |
| 2020 Presidential Election Index (Trump% - Biden%) | 0.307520076 | -0.344633775 |
| Social Capita | -0.31099029 | -0.407327764 |
| log(Cat/Dog) | -0.22165045 | -0.263981313 |

Table 7:

| topic | tweet |
|---|---|
| Mandatory Vaccination | well i think it's not ok to force some of us that don't want to the vaccine and it's also not ok to force someone to not take the vaccine either most of us don't like being force i think it's ok for us to make our own decision free will if some choose to take it or not |
| Mandatory Vaccination | Wish people would stop telling people which vaccine they should take let people decide themselves and give choice. People want vaccine but doesn't mean they want to take just anything someone tells them to take. |
| Mandatory Vaccination | All you're doing is taking our right our right to choose whether we want to take a vaccine whether we want to live or die you were taking all our rights away and it is not your right to take it away from us and don't tell me you're giving us the right to choose there is no choice |
| Mandatory Vaccination | the people act like you giving us the freedom to decide for ourselves is taking something from THEM. They act like they have to be mandated to take the vaccine to take it, instead of just having the freedom to choose to take it! People make YOUR decision not everyone's |
| Mandatory Vaccination | I also don't like how they are trying to take away our freedom free will if some of us choose to wear mask and get the vaccine or not wear mask to get vaccine i wish they respect our free will some of us don't like getting force to do things we don't wanna do |
| Mandatory Vaccination | Please have choices in life if they don't wanna take the vaccine that's their choice let people live the way they want to people that took the vaccine that was their choice stop the BS let people live their own life we should just get back to normal |
| Mandatory Vaccination | it's someone else's choice and freedom to decide if they want to take the vaccine or not so live your life however the fuck you want but don't tell people what to do with there's no one stopping you |
| Mandatory Vaccination | If you want the vaccine get it if you want to put a mask on put it on! But people have the right to choose if they want or do not want some thing not imposed by the government!!! |
| Mandatory Vaccination | And your trying to take away people's right not to get a vaccine. So the freedom of choice which you want for women, you want to take away from everyone else? Not sure how your being fair here |
| Mandatory Vaccination | "My point is, you don't have to force people to take it or use systematic censorship to force people to take it. ""My body my choice"" Let people make individual choices to take the vaccine. If the vaccines work why force people? Do you force people to eat? I need to eat to live" |
| Adverse reactions | Many vaccine-related deaths and serious side-effects have been reported in the UK and international media. As of 13 May 2021, there were 822,078 reported adverse reactions in the UK, including seizures, paralysis, blindness, strokes, blood clots and acute cardiac events |
| Adverse reactions | "*US where <10% adverse reactions reported to VAERS* ""(VAERS) revealed 294,801 reports of adverse events following COVID vaccines, including 5,165 deaths and 25,359 serious injuries between Dec. 14, 2020 and May 28, 2021.""" |
| Adverse reactions | <10 allergic reactions per 1 million doses 13 cases of blood clots per 1 million doses 11 cases of myocarditis per 1 million doses 1 case of swollen testicles per 1 million doses |
| Adverse reactions | Also vaccine-related deaths and serious side-effects have been reported in the UK and international media. As of 13 May 2021, there were 822,078 reported adverse reactions in the UK, including seizures, paralysis, blindness, strokes, blood clots etc, and 1178 deaths were reported |
| Adverse reactions | Please report reactions. Here is information about reporting an adverse reaction (VAERS link in the CHD article): ICAN reporting system CHD article on how to report a vaccine reaction UK reporting system |
| Adverse reactions | vaccines are causing serious adverse events and deaths. Interview the PA who reported vaccine injuries from her hospital. 680,000 reports of serious adverse events and thousands of deaths linked to the jab. |
| Adverse reactions | according to the under reported CDC 'VAERS' Vaccine Adverse Event Reporting System to November7 2021: 675,000 adverse events including 14,000 deaths, 18,000 disabilities 58,000 hospitalisations, 11,700 heart issues including heart attacks to cite a few: |
| Adverse reactions | adverse reactions - weekly report covering adverse reactions to approved vaccines 460 deaths & 243,612 injuries reported in |
| Adverse reactions | 685 deaths reported from adverse reactions to AZ vaccine. Over half a million adverse reaction reports. Link from UK Govt website. |
| Adverse reactions | VAERS, CDC Vaccine Adverse Event Reporting System logged most recent COVID injuries in America data. New figures report 20,000 vaccine related deaths. |

| topic | tweet |
|---|---|
| Constitutionality | To force vaccination is in violation of Nuremberg code UNESCO Universal Declaration on Bioethics and,Human Rights UN International Covenant of Civil and political rights UN Universal Declaration of Human Rights. No govt has the right to enforce vaccination |
| Constitutionality | so answer these basic questions do you support the following: Court packing? mandatory vaccination? mandatory milage tax? mandatory vaccination travel passport? Mandatory vaccination/testing on private business? These are the easier socialist policies being put forward answer plz |
| Constitutionality | Government requirements for private business to mandate vaccination of employees is a government mandate of human experimentation and a violation of the Nuremberg Code |
| Constitutionality | Joe Biden tells businesses to force vaccination for employees President Joe Biden encourage |
| Constitutionality | Vaccination passports are not for public health. Vaccination passports are for public control. |
| Constitutionality | In Texas, Gov. Greg Abbott has banned mask mandates, signed legislation that would deny state contracts or licenses to businesses that require proof of vaccination, and barred local governments from requiring the vaccine for any public agency or private institution. |
| Constitutionality | Show me where there is a federal and state requirement forcing private businesses to make vaccination mandatory for staff. |
| Constitutionality | Medical tyranny = mandated vaccination status required by the government to enter a private business, draconian lockdowns. |
| Constitutionality | Mandatory verification of vaccination status is a violation of HIPPA. Restaurants & Bars demanding your vaccination status are violating laws protecting your health information privacy. County officials pushing vaccination verification are in violation of HIPPA |
| Constitutionality | President Joe Biden ordered widespread vaccination mandates on Thursday, demanding that private businesses mandate vaccinations for their employees. doesn't give a damn about your constitutional civil rights or your health. Give Biden the boot! |
| Ineffectiveness | I got Facebooted from FB for 30 days for posting Vaxed people Can still get covid Can still spread covid Can still die from covid Can potentially die from the vaccine. Unvaxed people Can still get covid Can still spread covid Can still die from covid Cannot die from vaccine |
| Ineffectiveness | Twitter won't let u speak truth: Fact Vaxed people: Can still get covid. Can still spread covid. Can still die from covid. Can potentially die from the vaccine. Unvaxed people: Can still get covid. Can still spread covid. Can still die from covid. Cannot die from the vaccine. |
| Ineffectiveness | You mean the Vaccine that you get to not get Covid. Yet many people still get Covid and DIE from Covid even with the Vaccine? You mean that one? That's Exactly why I refuse to get it. The Polio shot,no one I knew got it after getting the shot. The Flu shot? Never get flu after it |
| Ineffectiveness | Man vaccinated against covid, who currently has covid, urges people not vaccinated against covid to get vaccinated against covid because the vaccine apparently works, even tho he still got covid and vaxed ppl are still dying from covid |
| Ineffectiveness | The only people getting Covid-19 are the ones getting the vaccine or their either dying after getting the vaccine. I never got the vaccine I dont believe In it. I also never got the flu shot and never had the flu. People who get the flu shot always get the flu nothing different |
| Ineffectiveness | Fact Vaxed people: Can still get covid. Can still spread covid. Can still die from covid. Can potentially die from the vaccine. Unvaxed people: Can still get covid. Can still spread covid. Can still die from covid. Cannot die from the vaccine. Its a no for my family |
| Ineffectiveness | "FACT Vaxxed People: Can still get Covid. Can still spread Covid. Can still die from Covid. Can potentially die from the vaccine. Unvaxxed People: Can still get Covid. Can still spread Covid. Can still die from Covid. Cannot die from the ""Vaccine""" |
| Ineffectiveness | You can die from the vaccine. You can get seriously injured from the vaccine. You can still get Covid with the vaccine. You can still die of Covid with the vaccine. You can still get terribly sick from Covid with the vaccine. You can still spread Covid with the vaccine. |
| Ineffectiveness | ive had the flu vaccine never got the flu or even sick from it, get the covid vaccine you still gonna get covid, be able to pass it, and some people still die from covid with the vaccine |
| Ineffectiveness | You can get the flu from a weakened virus in the vaccine which happens to me every time I get flu shot and get the flu 4 days later and am sick for 2-3 weeks and know others same thing. Daughter never gets sick..got the flu shot same time as another vaccine, 5 days later got flu |

| topic | tweet |
|---|---|
| Long Term Side Effects | There are short term,mid term and Long term side effects, long term side effects appear after 5, 7 or 10 years. Without knowing the long-term side effects no vaccine can be approved & it take at least 10 years to know Long term side effects. |
| Long Term Side Effects | mRNA vaccine is only experimental medicine which has zero long term safety data. mandatory experimental medicine without long term safety data policy is against humanity. |
| Long Term Side Effects | Those are "licensed"vaccines. This is a non-licensed emergency use authorized vaccine only. Meaning we have no long term data on side effects. Vaccines take five years for approval so they could study long-term side effects! We are the lab rats! |
| Long Term Side Effects | Vaccines have short and long term side effects, long term side effects appears after 3 to 10 years, for a vaccine to be called safe its necessary to have a long term safety data and long term safety data can only be obtained if the vaccine is tested for at least 10 years... (1/n) |
| Long Term Side Effects | You can argue for the science all you want, and the chances of long-term unexpected side effects are incredibly slim, but forcing children to get a vaccine using technology only approved for human use for a year, with no real-life long-term safety studies, is insane. |
| Long Term Side Effects | Cautiously yes, not with Emergency Use Authorisation before proper long term clinical trials. This vaccine is not even a year old and we have no idea about possible long term side effects yet. Give it to people at high risk with clear benefit but not healthy kids. |
| Long Term Side Effects | The idea that 12 - 17 year old kids can make an informed decision about a vaccine that uses new technology and has NO long term safety data and therefore potential long term risks is absurd. No one is making an informed decision as there is NO long term data! |
| Long Term Side Effects | Oh my God! What we don't know long-term is obviously the side effects of the vaccine itself. There is zero long-term safety data on these vaccines. Do you know of any long-term safety data which exists for mRNA vaccines? |
| Long Term Side Effects | Long-term effects Unknown, Emergency use of an Experimental Vaccine that is not FDA Approved or Fully tested. 15 years minimum to Roll out Fully tested Vaccine. Informed Consent for an Unknown Outcome is Impossible! |
| Long Term Side Effects | Informed Consent. Given this vaccine is a human live trial without long term testing Canadians must be given raw data and informed consent. Unknown long term side effects including organ health, reproductive health and cancer are unknown. Choices should be made individually. |
| Politics and Conspiracy Theories | We all know why Big Pharma will do anything to SUPPRESS Ivermectin !! The moment that Big Govt accepts cheap & accessible Ivermectin can be effective & would set us free quickly, it would stop Big Pharma making BIG $$$ Big Pharma JUST playing us for $$$ |
| Politics and Conspiracy Theories | Big Pharma & it would shed positive light on President Trump. HCQ costs a few cents per day & a new vaccine would make Big Pharma & Bill Gates richer. |
| Politics and Conspiracy Theories | Virus from China. Tests for virus from China Masks from China How much money is China making from their manufactured virus?? Every politician pushing the "vaccine" receives money from J&J, Modern-a & Pfizer among other big pharma places.. |
| Politics and Conspiracy Theories | We must investigate the corrupt CDC that lied to Americans. They hate President Trump so they do anything to destroy our economy. Dr Fauci and Dr Birx are in bed with evil Bill Gates foundation and big Pharma. Dr Fauci is pushing expensive therapy and bad vaccine to make billions |
| Politics and Conspiracy Theories | I'm tired of hearing 2 stories. I wish Trump would clarify it. Trump says Fauci, W.H.O, FDA, CDC, BIG PHARMA all corrupted. Bill Gates talked about DEPOPULATION. WHO CREATED THE VACCINE? If Bill Gates or Fauci are behind it like the world thinks, WHY IS TRUMP PUSHING THE VACCINE? |
| Politics and Conspiracy Theories | Big pharma sees profit potential with CoronaVirus being big money on vaccines. Big pharma also pays huge to MSM. Trump mentions cheap drugs that won't make big money. MSM hate him. Big Pharma offers drug reps to discredit Trump & cheap drugs. Vaccine stays relevant. MSM loves it |
| Politics and Conspiracy Theories | Biden Had No Plan and the Best Plan was Trump Plan the vaccine & Operation Warp speed Fauci & Big Pharma have no further science Dems doing dem things like the Blame Game |
| Politics and Conspiracy Theories | All these new strain of covid-19 all fake news spread by big pharma & their minions in the corrupt mainstream media! Every time you get a shot of vaccine it cost the government $40!now big pharma is talking about new vaccines for the new strain !they will make billion dollars |
| Politics and Conspiracy Theories | Create an issue, then sell the solution. $56B so far in vaccine profits. $350M to politicians from Big Pharma. Sales slow down ,create a bigger boogeyman & need for product. Americans are being lied to & sold out, Media & politicians bought by big Pharma. |
| Politics and Conspiracy Theories | We must open this country up now, Coronavirus is a big hoax. They must fire Dr Fauci & Dr Brix because they are evil in bed with big Pharma & Bill Gates foundation pushing horrible vaccines & drugs make mega billions. This vaccine will kill elderly & sick to control population |

| topic | tweet |
|---|---|
| Natural Immunity | Masks never effective for respiratory virus, asymptomatic transmission never driver for respiratory virus, natural immunity > vaccine immunity, vaccine can't prevent transmission or infection, viral load in vax same as non vax, CDC admit no transmission from previously infected. |
| Natural Immunity | Vaccine can give a higher antibody count than natural immunity, but natural immunity gives immune cell memory which results is superior immunity than vaccinated immunity. Natural immunity also recognizes variants. |
| Natural Immunity | Those unvax'd who recover from infection have superior immunity to reinfection vs. vax'd who were never infected. Natural immunity is broader and more durable than vax immunity. Mass vaccination w/leaky vax in the middle of a pandemic is insane. Follow the science. |
| Natural Immunity | The vaccine does not prevent infection or transmission. The vaccine does not prevent infection or transmission. The vaccine does not prevent infection or transmission. The vaccine does not prevent infection or transmission. The vaccine does not prevent infection or transmission. |
| Natural Immunity | Herd immunity has historically been based on natural infection. Vaccination could help provide herd immunity IF it stops transmission. But it doesn't. So no chance of herd immunity from the vaccine. Infected-recovered rarely re-catch (spread) the virus so herd immunity develops. |
| Natural Immunity | Have you considered natural immunity far more robust than vaccine immunity, vaccines do not prevent transmission or infection, viral load in vaccinated same as non vax, masks never effective for a respiratory virus, asymptomatic transmission never a driver for respiratory virus ? |
| Natural Immunity | "-Vaccine ""immunity"" is less durable than natural immunity (memory B and T cells) -True, but Natural Immunity is already more effective than double Vax -The vaccines have been proven to be causing selection and producing variants, not natural infection - this is completely false" |
| Natural Immunity | The viruses mutates every time it replicates, the idea is to prevent escape mutants (super viruses) natural immunity prevents this. vaccination actively encourages super viruses. Yes the virus stops at natural immunity, when herd immunity is reached, . |
| Natural Immunity | You don't need both and natural immunity is far superior to the vaccine immunity in preventing reinfection and at limiting symptoms in those who are reinfected. All that means is that natural immunity is better to stop the spread and create herd immunity. |
| Natural Immunity | Vaccine does not prevent you from having the virus again and there is no evidence the immune response mounted by the vaccine is better than natural immunity. In fact for most, natural immunity would give rise to a stronger immune response and thus provide better protection. |
| Others | i hav entirrely opted out of your [all of your |
| Others | The is The is engineered The is engineered The is engineered The is engineered The are engineered Your is engineered The is engineered IS ENGINEERED |
| Others | $HTBX Understanding of Tcells for $GNUS $NBY $KODK $IBIO $GAXY |
| Others | Evidence that the are not as efficacious as advertised by |
| Others | This is what I am referencing my "opinion" of the vaccine. |
| Others | Hilarious how all these manufacturers touted themselves as a "vaccine" originally . |
| Others | He is dreaming Boris, I will not have any "vaccine " |
| Others | And it will about as effective as the flu vaccine. $$$$$$$$$$$$$$$$$$$$$$$$$$$$$$... |
| Others | Phuck that vaccine I'm not Turing into no zombie |
| Others | "the vaccine is ushering in the mark of the beast," |
